# Exploring the potential benefits and challenges of artificial intelligence for research funding organisations: a scoping review

**DOI:** 10.1101/2024.09.26.24314280

**Authors:** Amanda Jane Blatch-Jones, Hazel Church, Ksenia Crane

## Abstract

**Background:** Artificial Intelligence (AI) is at the forefront of today’s technological revolution, enhancing efficiency in many organisations and sectors. However, in some research environments, its adoption is tempered by the risks AI poses to data protection, ethics, and research integrity. For research funding organisations (RFOs), although there is interest in the application of AI to boost productivity, there is also uncertainty around AI’s utility and its safe integration into organisational systems and processes. The scoping review explored: *‘What does the evidence say about the current and emerging use of AI?’; ‘What are the potential benefits of AI for RFOs?’* and *‘What are the considerations and risks of AI for RFOs?’*

**Methods:** A scoping review was undertaken with no study, language, or field limits. Due to the rapidly evolving AI field, searches were limited to the last three years (2022-2024). Four databases were searched for academic and grey literature in February 2024 (including 13 funding and professional research organisation websites). A classification framework captured the utility and potential, and considerations and risks of AI for RFOs.

**Results:** 122 eligible articles revealed that current and emerging AI solutions could potentially benefit RFOs by enhancing data processes, administration, research insights, operational management, and strategic decision-making. These solutions ranged from AI algorithms to data management platforms, frameworks, guidelines, and business models. However, several considerations and risks need to be addressed before RFOs can successfully integrate AI (e.g., improving data quality, regulating ethical use, data science training).

**Conclusion:** While RFOs could potentially benefit from a breadth of AI-driven solutions to improve operations, decision-making and data management, there is a need to assess organisational ‘AI readiness’. Although technological advances could be the solution there is a need to address AI accountability, governance and ethics, address societal impact, and the risks to the research funding landscape.

## INTRODUCTION

Digital transformation is a cornerstone of innovation and progress. Sectors from healthcare to industry now operate on vast volumes of (‘big’) data and leverage it to stay competitive, deliver services, and meet societal needs. At the forefront of this technological revolution (“Industry 4.0”(^1–3^)) is innovation in data science and artificial intelligence (AI). AI is a collective term for technologies that enable ‘machines’ to emulate various complex human activities(^4^), and their unique capabilities (autonomy, adaptivity) give them almost limitless societal value (e.g., as ‘productivity’ tools, or for making predictions in data).(^5^)

The unique capabilities of AI also lend to its diverse applications, particularly for the purposes of research. large language models (LLMs), like ChatGPT, have for instance enabled students and staff to develop and enhance research skills (e.g., academic writing), while machine learning (ML) algorithms have been used to enhance and/or automate routine research tasks, such as data analysis.(^6–9^) The impact of these technological advancements on education and research has been transformative, bolstering wider interest, investments, and public initiatives in AI/ML technologies.(^10–14^) The use of AI/ML in research is rapidly growing, with more tools becoming available, and the user base broadening from academic institutions to research funding organisations (RFOs) and government bodies that commission and fund societal research. However, the perceived benefits of implementing AI in different research contexts have also been tempered by growing concerns of potential impacts on ethics and research integrity.(^15–19^) With the rapid advancements in AI technology outpacing regulatory efforts, the potential for misuse (e.g., plagiarism) or excessive reliance on AI tools in research is becoming a growing concern.(^20–22^) The apprehension this has created among organisations, stakeholders and users in research has triggered a global response to regulate and ensure the responsible use of AI.(^23–28^) Through initiatives such as Responsible AI UK (https://rai.ac.uk/), governments and RFOs are introducing more evidence-based AI practices, guidelines and human oversight into AI-driven research activities.(^24, 29, 30^)

For RFOs, it is crucial to explore the application of AI/ML technologies, which although may have the potential to significantly reduce administrative research burden, could also bring substantial risk to public, organisational and funding activities.(^31–33^) However, efforts have been constrained by a lack of relevant evidence, which has so far predominantly focused on the use of academic applications of AI such as in learning environments or for grant writing and peer review.(^17, 34–36^) For publicly funded research, it is becoming increasingly burdensome due to stricter compliance and the volumes of data now needed to monitor and report on the outputs, outcomes and impacts of funded research.(^31–33^) As a result, RFOs find themselves managing growing research portfolios while coping with increasing research bureaucracy and data management issues created by the use of multiple digital systems.(^33, 36^) While there is a potential role for AI in reducing research bureaucracy (e.g., supporting risk assessment, automating data entry), many RFOs feel the need to tread carefully, due to concerns around issues such as data protection and privacy that creates understandable hesitancy around AI adoption.(^37^) There is also uncertainty around AI’s utility, the safety of commercial AI solutions, the value to organisational efficiency, and the risks of AI integration into established RFO enterprise processes.(^38, 39^) As RFOs assess their capabilities and needs in this space, there is a need to review current published literature and be guided by latest evidence to gain insight (from across sectors and settings) on what AI is, what it is not, what it can be used for, and how it could be implemented.

To address the above, and inform RFOs with a better understanding of the potential utility of AI within an organisational context (e.g., data management, research, administrative and operational efficiency) a scoping review was conducted to explore: *‘What does the evidence say about the current and emerging use of AI?’; ‘What are the potential benefits of AI for RFOs?’* and *‘What are the considerations and risks of AI for RFOs?’*

## METHODS

A scoping review approach was chosen due to the breadth and complexity of the topic and the available research in terms of the source, type, and setting. Scoping reviews help address priority questions in research, clarify concepts, and provide background information or frameworks in preparation for a systematic review (where applicable). Scoping reviews typically seek to identify evidence gaps or scope a body of literature, rather than describing experiences or what interventions work for particular individuals.(^40^) Therefore, scoping review methodology does not judge the quality of the evidence but rather maps the evidence.

The JBI (Joanna Briggs Institute) scoping review framework was used to guide the development of the review, including the searches, data analysis and reporting (https://jbi.global/scoping-review-network).(^41^)

To ensure the evidence was mapped according to areas of relevance for RFOs, a classification framework was developed (^6, 7^) to capture the 1) utility and potential and 2) considerations and risks associated with the use of AI for RFOs (see **Fig 1**).

**Fig 1.**
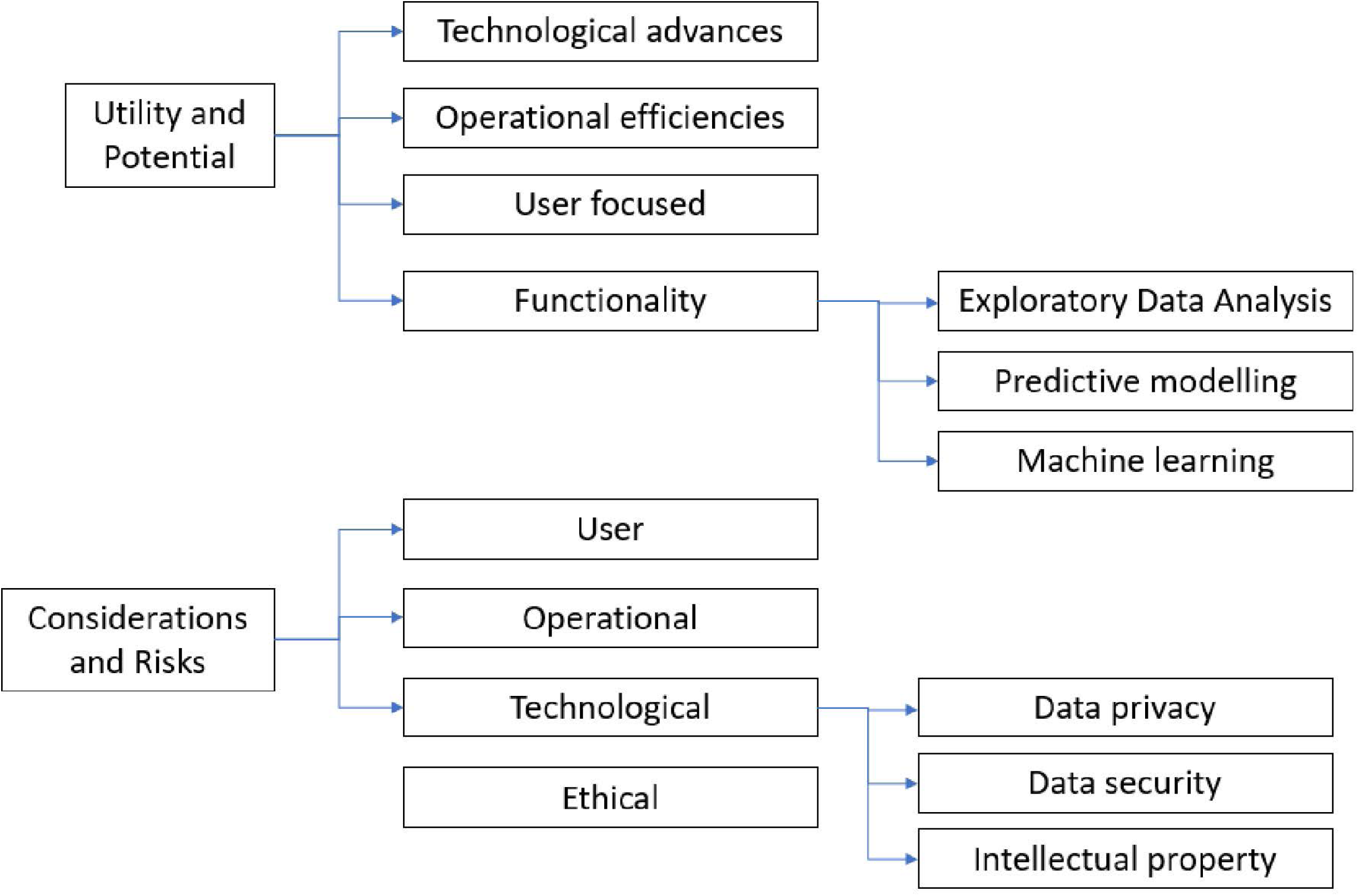
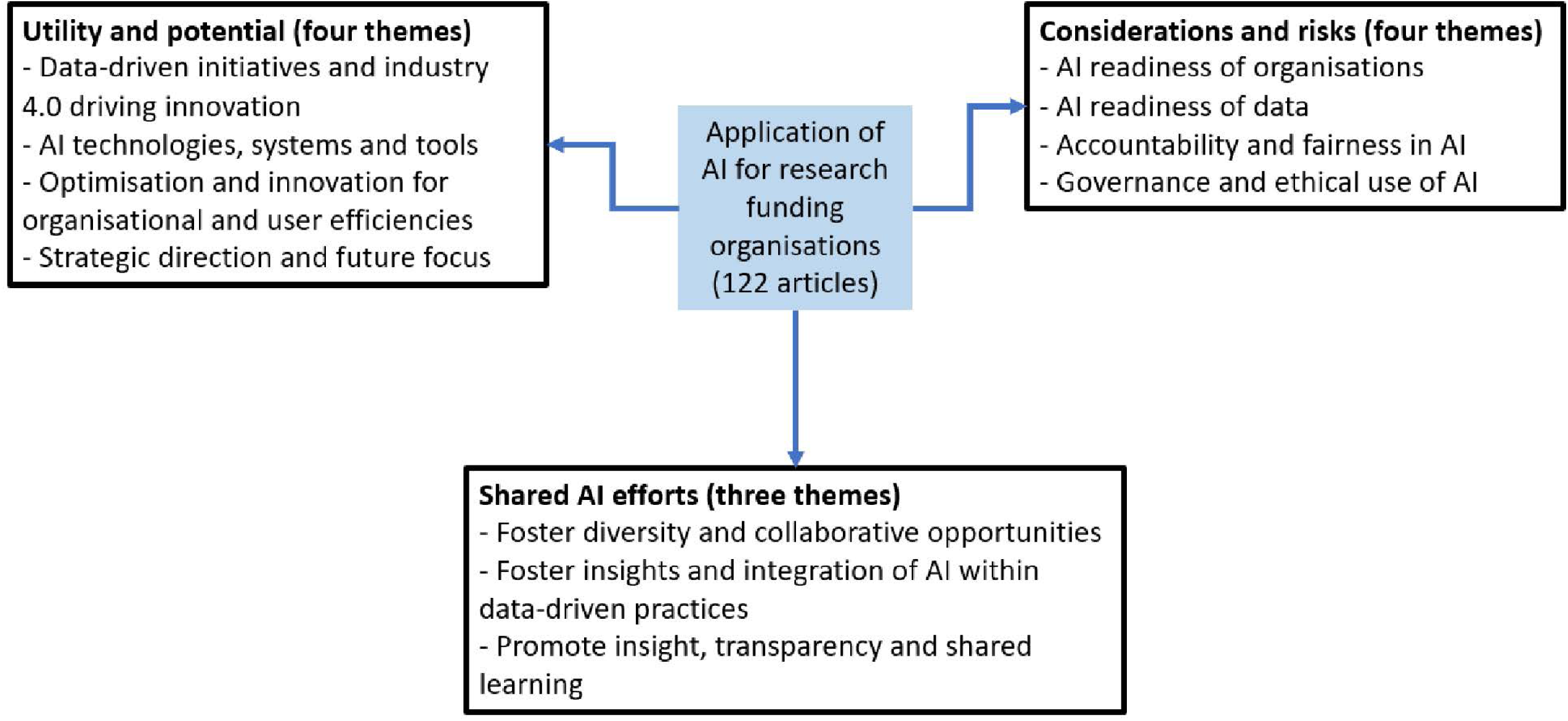
Utility, potential, considerations and risks of AI for research funding organisations classification framework to summarise the evidence

### Eligibility criteria

There is no universally accepted definition of ‘artificial intelligence’, so for the purpose of this review it was important to clarify what we meant by ‘AI’. Below are some previous explanations of AI that provided context for the review, including how we developed the eligibility criteria.

- ‘Artificial Intelligence’ is an umbrella term for an array of technologies possessing the unique capabilities of ‘adaptability’ and ‘autonomy’. This means that AI systems are trained by humans to infer connections and patterns in pre-defined data but can also generate new data and use it to make independent decisions.(^42–44^)
- AI can be simply defined as “a technology that enables machines to imitate various complex human skills” but others may also use more precise definitions, referring to specific capabilities and AI classification taxonomies.(^45^)
- Newman *et al.* (2022) defined AI as a “description of various computer operations designed to replicate human intelligence…It is a component of a greater set of computing technologies that includes automated decision making, advanced algorithms, and data processing at large scale.”(^46^)
- Today’s AI models can be broadly categorised into ‘predictive AI’ (analysing patterns in predefined datasets to make predictions on new data, for example ML) and ‘generative AI’ (generating new data on the basis of predictions from patterns in datasets, for example LLMs). (^17, 47, 48^)

#### Context

The context included UK, European and international settings (e.g., sectors, fields or disciplines) (based on the inclusion criteria) and were included to gain as much information as possible on potentially relevant AI applications.

#### Participants

Research funding and performing organisations, private sector business and enterprise, and public administrations were included as part of the AI application context.

#### Inclusion criteria

Articles were included if they were written in the English language (or could be transcribed to English), available in full text, and focused on the application and implementation of AI (rather than the development of algorithms). To be relevant to RFOs, these AI applications had to apply primarily to text data (^16^) (rather than images) used for/in organisational processes in an institutional, enterprise or business context (comprising specific human tasks and operational processes). We defined ‘organisational processes’ to include (but not be limited to): research management and administration; business intelligence; compliance and risk management; data management and analytics; and decision-making.

With regards to capturing relevant AI solutions:

- All included articles had to comply with the classification framework(^6, 7^), whereby the evidence had to inform on either the 1) utility and potential of AI (e.g., technological advances, operational efficiencies, user focus, and functionality), and/or 2) the considerations and risks of AI (e.g., user, operational, technological (including data privacy and security, and intellectual property), and ethical), or preferably both.
- All included articles on AI solutions had to explain where, how and why these solutions have been, or could potentially be, applied to enhance organisational processes (e.g., for administrative efficiency and effectiveness).
- Relevant articles could include any type of AI system, model, or tool, whether already available and/or marketed (e.g., OpenAI’s ChatGPT, Cloud Solutions, Amazon Web Services) or in the research and development/piloting stage (e.g., predictive models, ML).
- Relevant articles could also discuss infrastructures that support the integration and implementation of AI by organisations, such as data management and storage solutions (e.g., data lakes, data warehouses) or Information and Communication Technology (ICT) solutions (e.g., Network solutions, PowerSwitch).
- Relevant articles could refer to other types of solutions relevant to AI implementation (but not software or tools), such as regulative laws or guidelines, business models and frameworks, recommendations, and standards (e.g., data standards or data management plans) that could support organisational integration of AI.

#### Exclusion criteria

We excluded any articles that were retracted or provided limited information on AI applications, as defined in the inclusion criteria. News announcements related to company partnerships or the marketing of new products were excluded, as well as articles on AI applications not relevant to an organisational context (e.g., military or medical AI applications). The following is a non-exhaustive list of AI applications that were excluded from the scoping review: Drone technology; 3D printing; environmental and agricultural (e.g., fire safety, water reserves, farming, drainage, space); satellite data and weather monitoring; facial and voice recognition; metaverse and mobile networking (5G and 6G); estimation and stimulation modelling (e.g., urban flood, mining, energy grids); medical education; healthcare intervention; medical devices and diagnostics.

### Source types

All article types and study designs were included in the review to maximise coverage due to the diverse nature of the use and adoption of AI applications across different settings and sectors (which could be relevant to RFOs). We therefore included:

#### Academic type articles

Peer reviewed journal articles; commentaries; editorials; opinion letters; perspectives; and preprints

#### Grey literature

Policy documents; reports; blogs; educational articles; guidance; and newsletters (e.g., AI News, AI Magazine).

#### Study designs

Randomised controlled trials; non-randomised controlled trials; before and after studies and interrupted time-series studies; analytical observational studies (including prospective and retrospective cohort studies); case-control studies; analytical cross-sectional studies; descriptive observational study designs (including case series); individual case reports; and descriptive cross-sectional studies. Qualitative studies were also considered that focused on qualitative data including, but not limited to, phenomenology, grounded theory, ethnography, qualitative description, action research and feminist research. In addition, systematic reviews that met the inclusion criteria were considered, depending on the research question.

### Search strategy

Searches were conducted on 1 February 2024, using Web of Science and ProQuest databases to search for published journal articles, and Overton and Google Scholar were used to search for grey literature and government or RFO documents. The search strategy was iteratively developed and adapted to each database, with keywords and search terms identified from pilot searches in Web of Science, and manual searches in Connected Papers (https://www.connectedpapers.com/), to inform the final search strategy (**See S1 Appendix: Search terms and keywords** and **S1 Table: Search strategies**). The final search strategy was reviewed and approved by a librarian information specialist at the University of Southampton.

Searches were not limited by study type or design, source of publication, country, or language of publication. Initially, no limits were placed on year of publication; however, during initial screening of the title and abstract, it was noted that those published before 2022 were consistently out of scope and did not meet inclusion criteria for the current review (e.g., articles were too conceptual or focused on algorithm development only). Subsequent searches were limited to articles published in the last three years only (2022–2024) and this was reflected in the search strategy and screening process in accordance with PRISMA (https://www.prisma-statement.org) (**see Fig 2** and **S2 Appendix: PRISMA checklist**). Articles deemed relevant by title and abstract were then screened by reading the full texts of the articles.

**Fig 2.**
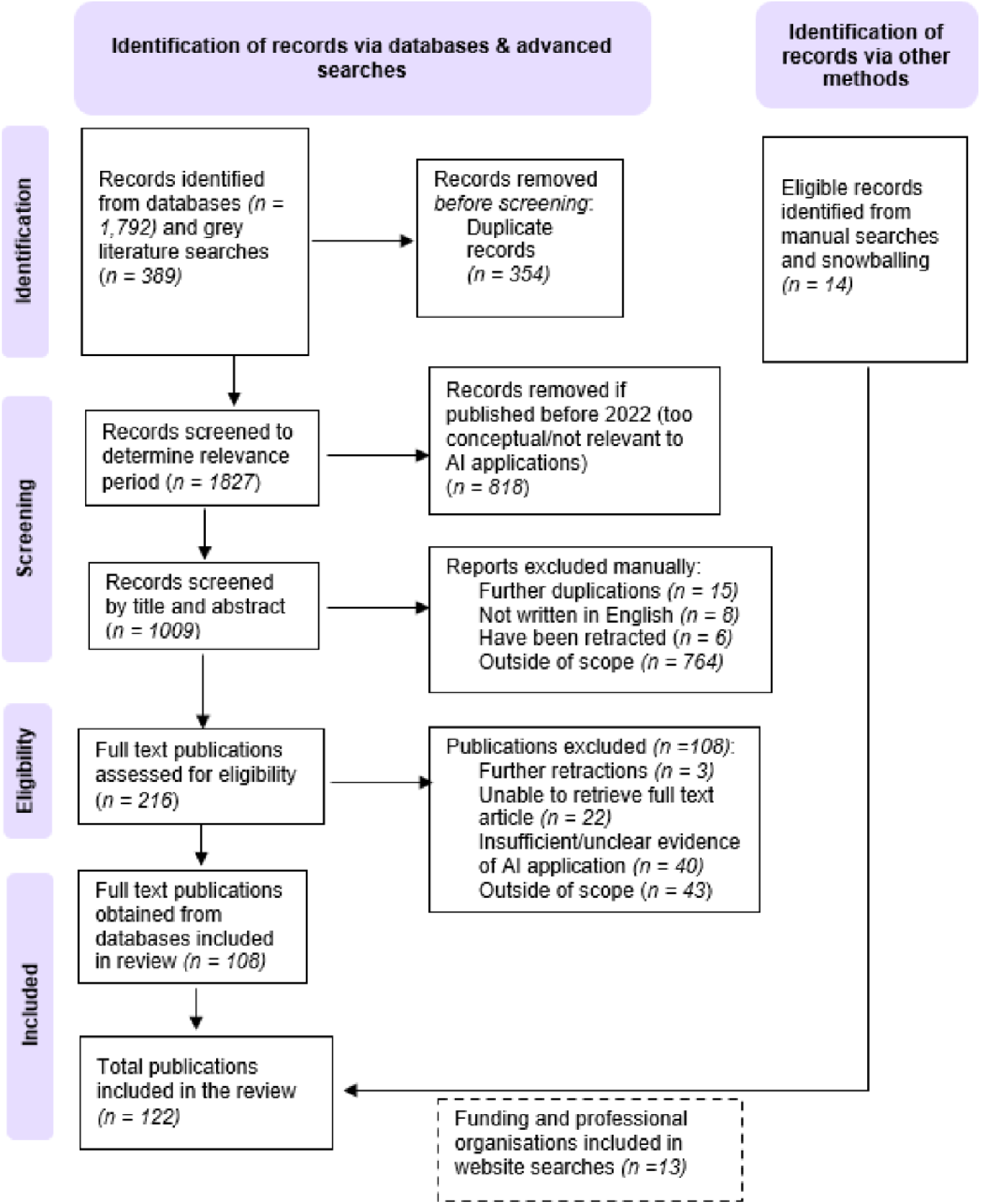
PRISMA flow diagram of the search strategy used to identify articles, research funding and professional organisations websites and the results of the searches.

Manual searches were also conducted on 13 websites of research funding and professional organisations (e.g., research councils, charities and professional/regulatory bodies) to identify announcements, funding opportunities, or other activities relevant to AI. This was done by typing the name of each organisation into the Google search engine and navigating to relevant webpages. The organisations were chosen by purposive sampling, following discussions with National Institute for Health and Care Research (NIHR) Coordinating Centre staff, and aimed to be representative of positions, practices and developments in big data and AI across the research funding landscape.

### Data extraction

The articles identified through the databases and manual searches were exported with citation, titles, and abstracts into Endnote 20 (Clarivate, UK). Duplicates were removed using the EndNote duplication function and then manually checked for consistency. To test eligibility criteria and ascertain agreement, 20 articles were assessed in a pilot screen by two authors (ABJ and KC). All remaining articles were screened by title and abstract against the inclusion criteria. Relevant articles were then checked for availability of full texts and the full texts were then screened by both authors to check for eligibility. Any disagreements between authors regarding the decision to include or exclude were resolved through discussion until consensus was reached.

After the final screening, ABJ and KC extracted data from the article’s full texts into a Microsoft Excel data extraction spreadsheet developed by the authors specifically for this review and was piloted to make sure the relevant information was extracted to address the three research questions and the classification framework. The spreadsheet included the following sections for extraction:

- Demographics: article contributors; journal; year; geographical location/s; source type; study type (if relevant)
- Relevant information on the AI application/s: organisational or sector settings; area/s of AI application; aim; content and purpose of the article; summary of any current issues mentioned and AI solutions (if applicable)
- A classification framework was used to capture the evidence under two areas: utility and potential of AI (e.g., technological advances, operational efficiencies, user focus, and functionality) and considerations and risks of AI (e.g., user, operational, technological (including data privacy and security, and intellectual property), and ethical).(^6, 7^) (see **Fig 1**)

Where applicable, further exclusions were made during the full data extraction stage, for example, where the same article was included multiple times due to different source types (i.e., duplication of information). In such instances, a peer reviewed journal article took precedence (and was included) over a preprint manuscript and a conference proceeding, and a preprint manuscript took precedence over a conference proceeding. For peer reviewed articles, if the same article was published in two journals, only one was included. Any articles written in languages other than English, or lacking an English translation, were excluded from full extraction.

All extracted data were used to inform summary tables characterising the included articles and drawing out the key concepts to address the three research questions (refer to Results section for **Tables 2, 3, and 4**). As this was a scoping review, we did not conduct a risk of bias or quality assessment, and all evidence was mapped or categorised to either pre-defined terms and concepts or to those developed and agreed by the authors during the data extraction stage. The results of the searches and study inclusion process are reported in full, in accordance with the Preferred Reporting Items for Systematic Reviews and Meta-analysis extension for scoping reviews (PRISMA-ScR) (see **Fig 2**).

**Table 1:**
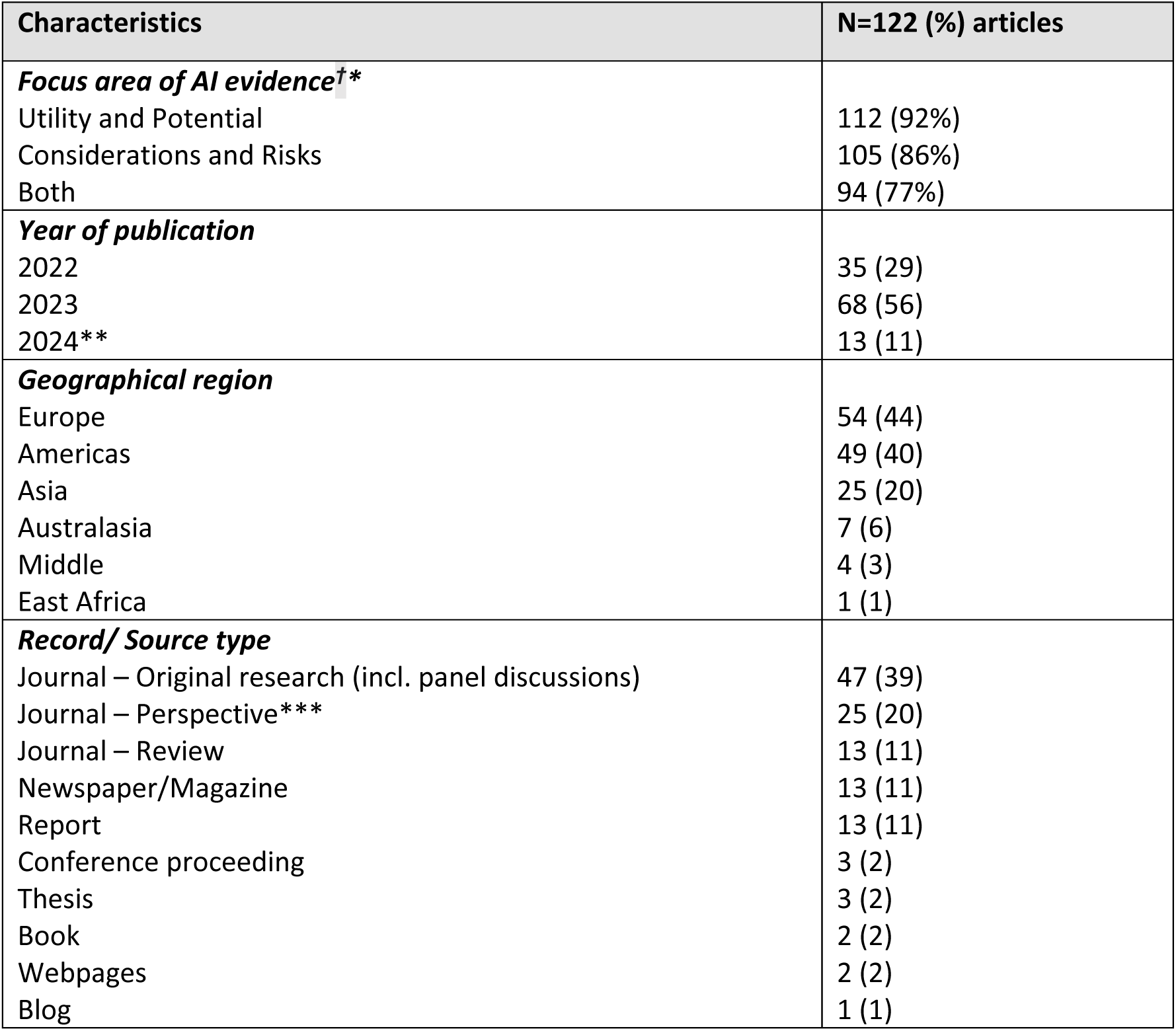
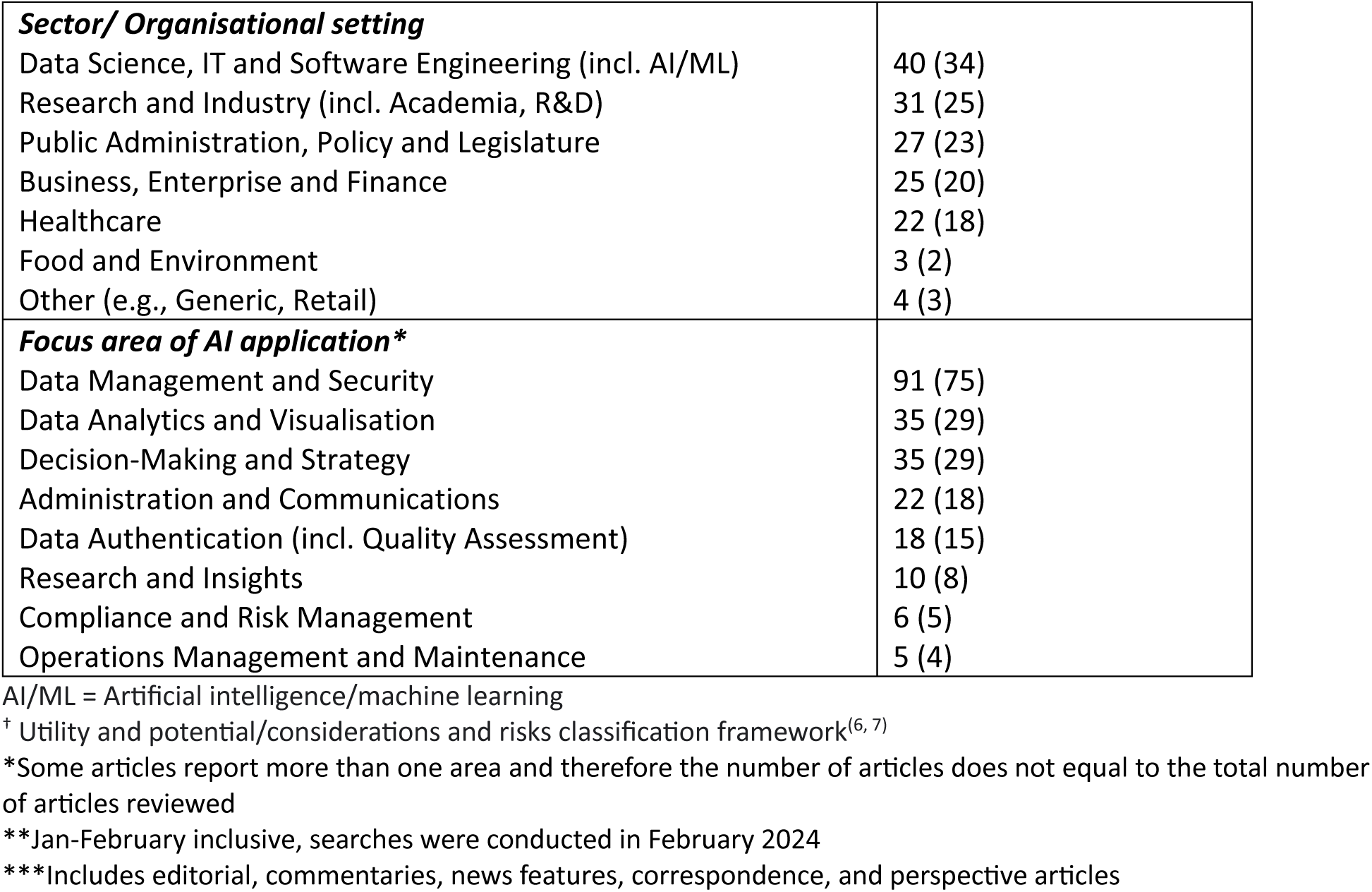
Characteristics of the included articles.

| Characteristics | N=122 (%) articles |
| --- | --- |
| ***Focus area of AI evidence<sup>†\*</sup>*** |  |
| Utility and Potential | 112 (92%) |
| Considerations and Risks | 105 (86%) |
| Both | 94 (77%) |
| ***Year of publication*** |  |
| 2022 | 35 (29) |
| 2023 | 68 (56) |
| 2024** | 13 (11) |
| ***Geographical region*** |  |
| Europe | 54 (44) |
| Americas | 49 (40) |
| Asia | 25 (20) |
| Australasia | 7 (6) |
| Middle | 4 (3) |
| East Africa | 1 (1) |
| ***Record/ Source type*** |  |
| Journal – Original research (incl. panel discussions) | 47 (39) |
| Journal – Perspective*** | 25 (20) |
| Journal – Review | 13 (11) |
| Newspaper/Magazine | 13 (11) |
| Report | 13 (11) |
| Conference proceeding | 3 (2) |
| Thesis | 3 (2) |
| Book | 2 (2) |
| Webpages | 2 (2) |
| Blog | 1 (1) |
| **Sector/ Organisational setting** |  |
| Data Science, IT and Software Engineering (incl. AI/ML) | 40 (34) |
| Research and Industry (incl. Academia, R&D) | 31 (25) |
| Public Administration, Policy and Legislature | 27 (23) |
| Business, Enterprise and Finance | 25 (20) |
| Healthcare | 22 (18) |
| Food and Environment | 3 (2) |
| Other (e.g., Generic, Retail) | 4 (3) |
| **Focus area of AI application\*** |  |
| Data Management and Security | 91 (75) |
| Data Analytics and Visualisation | 35 (29) |
| Decision-Making and Strategy | 35 (29) |
| Administration and Communications | 22 (18) |
| Data Authentication (incl. Quality Assessment) | 18 (15) |
| Research and Insights | 10 (8) |
| Compliance and Risk Management | 6 (5) |
| Operations Management and Maintenance | 5 (4) |

AI/ML = Artificial intelligence/machine learning
<sup>†</sup> Utility and potential/considerations and risks classification framework<sup>(6, 7)</sup>
\*Some articles report more than one area and therefore the number of articles does not equal to the total number of articles reviewed
\*\*Jan-February inclusive, searches were conducted in February 2024
\*\*\*Includes editorial, commentaries, news features, correspondence, and perspective articles

**Table 2:**
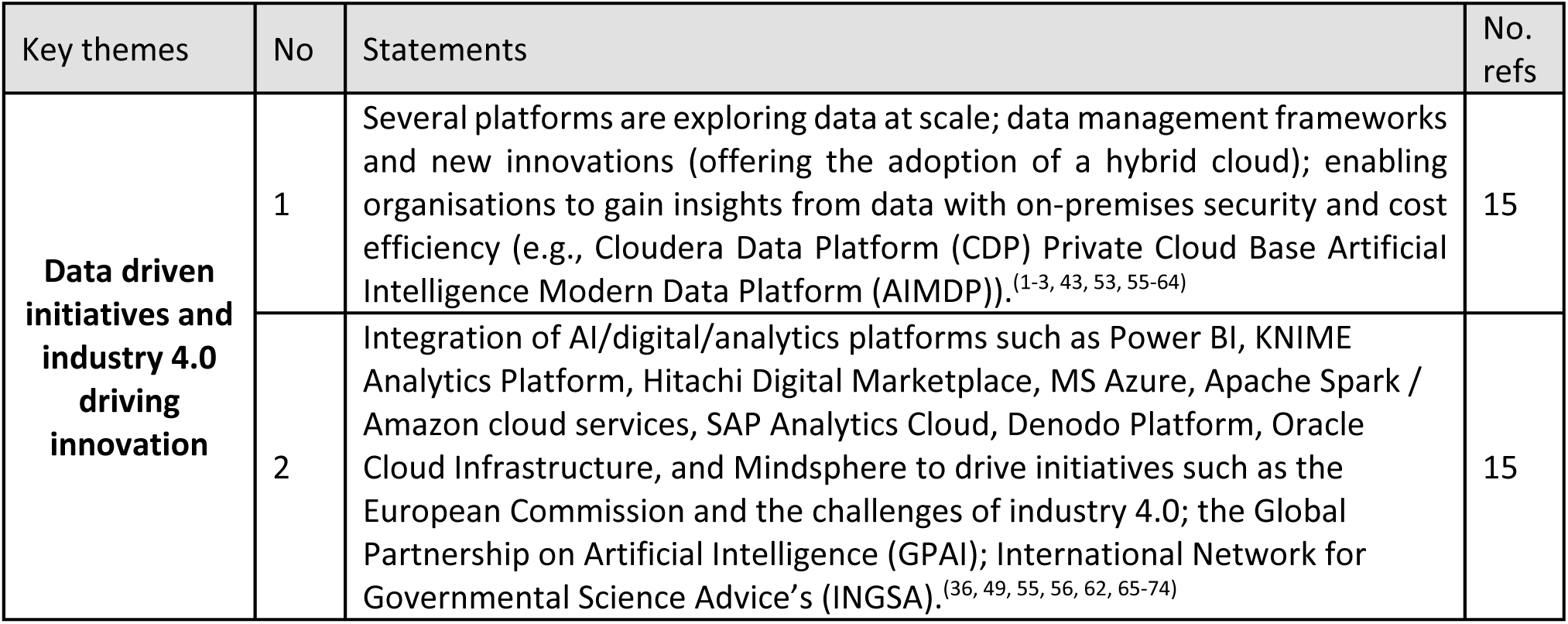
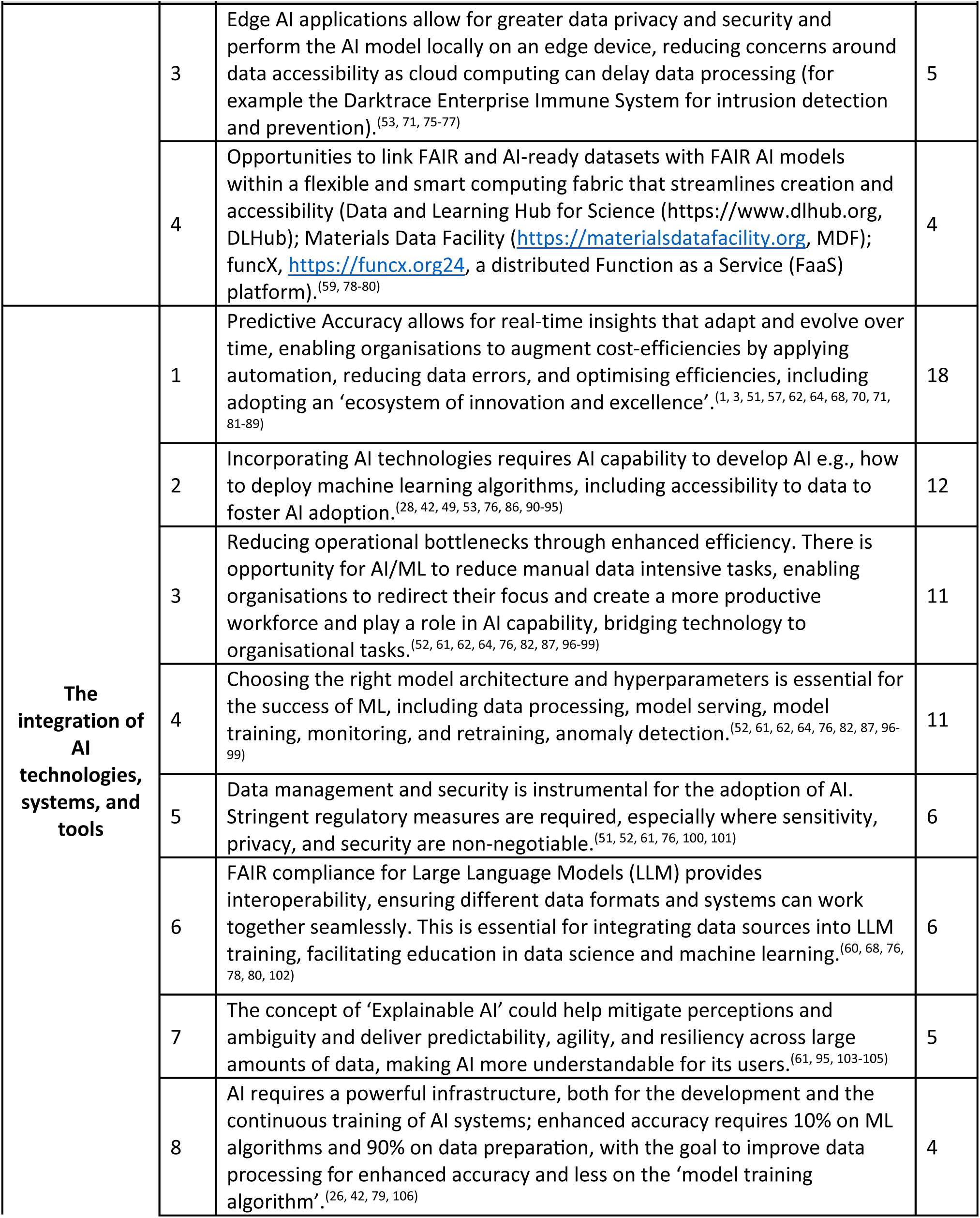
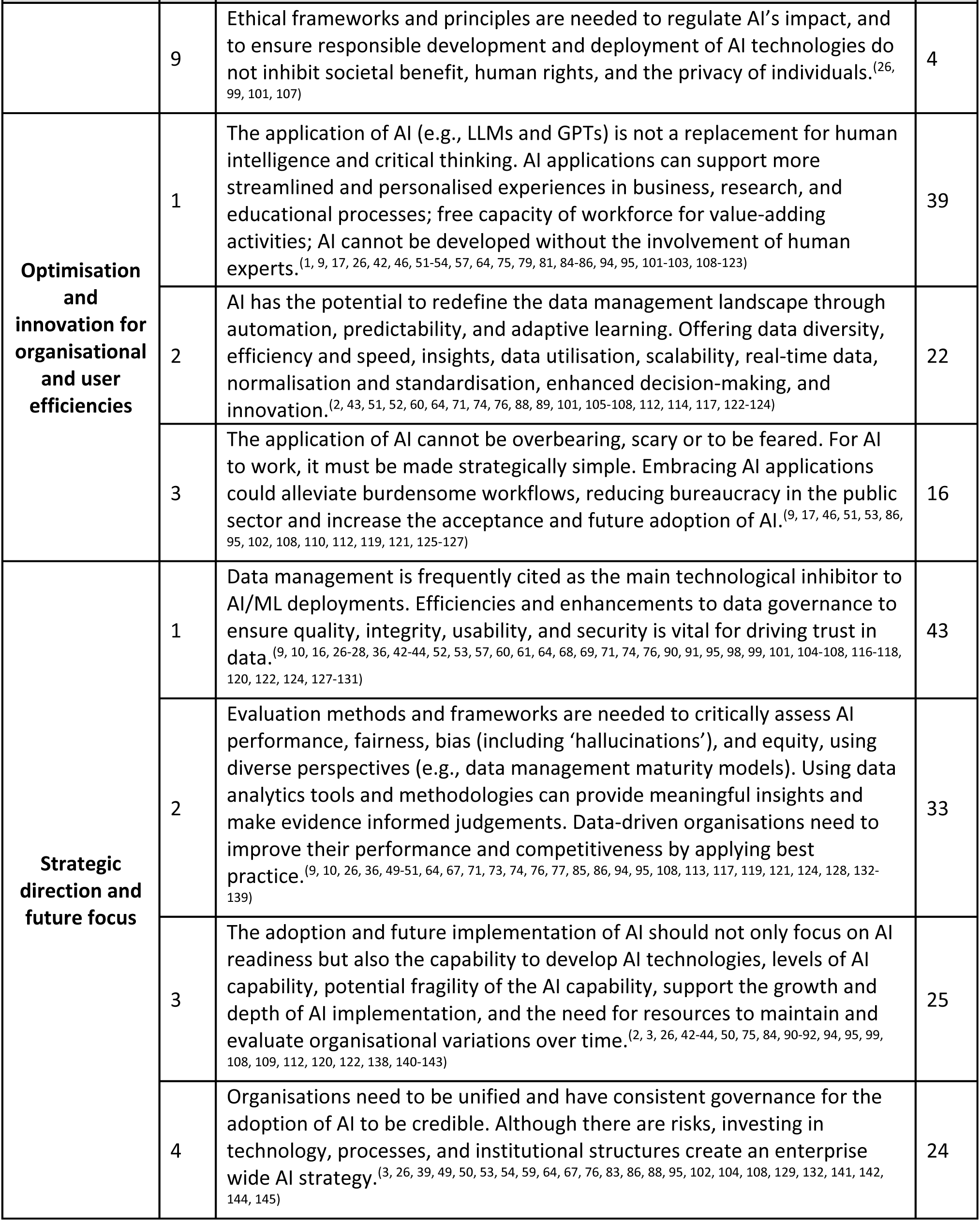
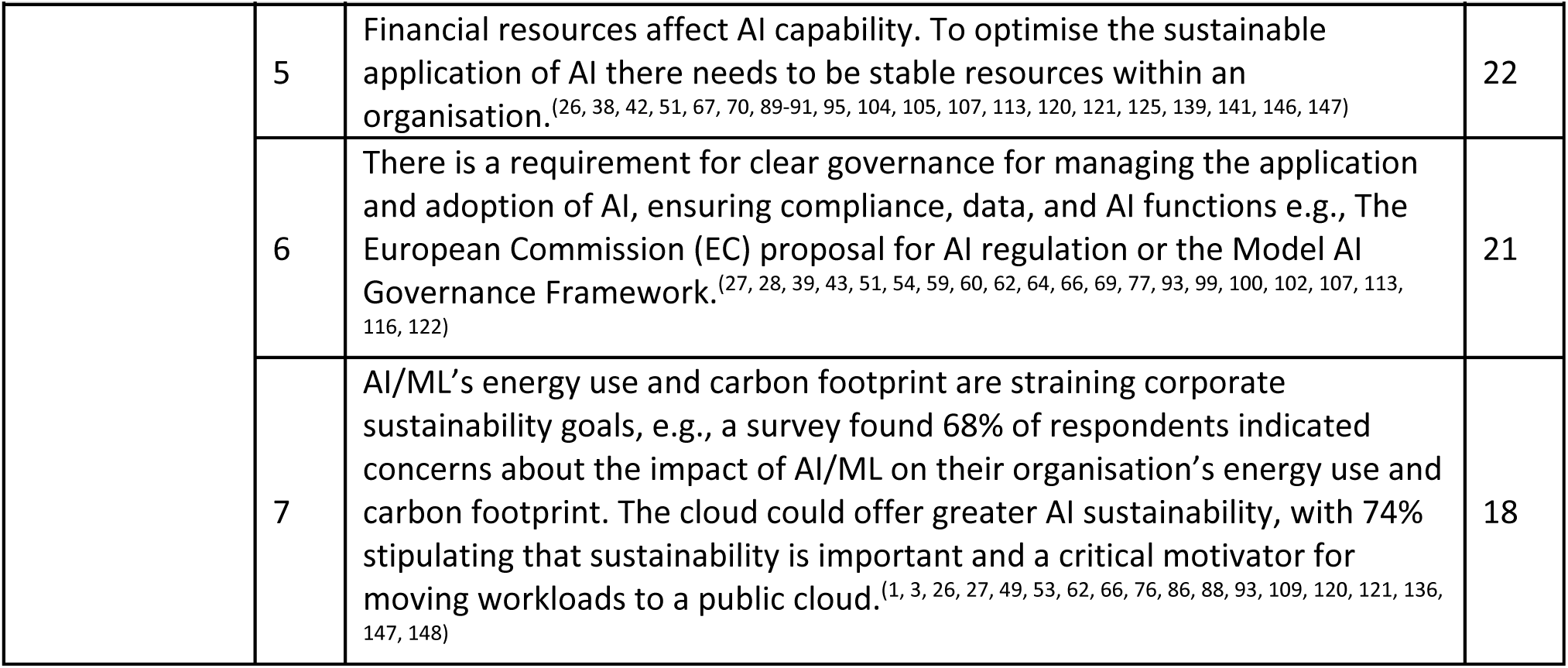
Summary statements from the included articles on the utility and potential of AI for research funding organisations.

**Table 3:** Summary of statements from the included articles on the considerations and risks of AI for research funding organisations.

| Key themes | No | Statements | No. refs |
| --- | --- | --- | --- |
| **Support the AI readiness of organisations** | 1 | Organisations must ensure they can cope with the unique challenges of AI, have the required infrastructure in place and establish AI model lifecycle capabilities.<sup>(26, 39, 50, 53, 61, 66, 75, 76, 78-80, 86, 94, 99, 101, 104, 107, 109, 113, 116, 119, 121, 123, 127, 131, 142, 147, 151)</sup> | 28 |
|  | 2 | Data leaders and research office staff need to acknowledge the need for increased investment into data management and AI solutions in research.<sup>(3, 10, 16, 26, 27, 39, 49, 50, 53-55, 76-78, 86, 88, 92-95, 100, 105, 120, 124, 145, 147)</sup> | 26 |
|  | 3 | There are concerns about AI model maturity, compatibility and interoperability, the expectations of users, and the costs of equipment and training.<sup>(1, 3, 16, 28, 50, 54, 59-62, 64, 68, 71, 74, 80, 81, 95, 98, 132, 139, 145)</sup> | 21 |
|  | 4 | Embracing new data norms and tools will require workforce adjustments to include data scientists, librarians or archivists, data management experts and AI/ML researchers.<sup>(3, 50, 54, 63, 74, 79, 92, 95, 100, 106, 109, 117, 144, 151)</sup> | 14 |
|  | 5 | Organisations already using AI report similar barriers around AI knowledge and digital skills, data quality and data privacy and protection.<sup>(26, 28, 53, 82, 91, 92, 95, 97, 98, 108, 121)</sup> | 11 |
| **Support the AI readiness of data** | 1 | Data management is the most frequently cited inhibitor to AI adoption and data integrity is seen as the top risk.<sup>(1, 9, 16, 26, 37, 43, 44, 49, 51, 52, 55, 59, 63, 64, 68, 69, 74, 76, 77, 81, 84, 86, 87, 89, 91, 97, 99-101, 104-106, 108, 112, 114, 116, 119, 121-123, 134, 136, 139, 141, 143, 144, 151, 152)</sup> | 48 |
|  | 2 | The need for better data management in research is increasingly recognised, for both RFOs, researchers and government bodies.<sup>(1, 9, 43, 44, 46, 49, 54, 63, 64, 74, 81, 84, 87, 88, 92, 98, 102, 104-106, 112, 119-121, 123, 126, 127, 136, 139, 141, 145)</sup> | 31 |
|  | 3 | A data-centric approach to AI development should focus on tools for data management, visualisation, and error detection, practical FAIR definitions for software and workflows, and frameworks for quantifying the FAIRness of AI models. <sup>(50, 52, 59, 60, 74, 78-80, 87, 89, 99, 101, 105, 106, 115, 126, 131, 134, 139)</sup> | 19 |
|  | 4 | Research data needs to be FAIR (findable, accessible, interoperable, and reusable) to ensure accuracy and performance of AI models. This requires changes to research practices, guidance, examples, and consistent metrics for evaluating data and model FAIRification.<sup>(50, 59, 60, 74, 78-80, 87, 99, 101, 105, 126, 131, 134)</sup> | 14 |
|  | 5 | Data must be cleaned and verified before it can be fed into algorithms to prevent bias and errors. This stage requires the most effort, but its importance is underestimated (for instance, compared to model development).<sup>(9, 52, 64, 78, 87, 104, 106, 116, 120, 126, 151)</sup> | 11 |
| **Support accountability and fairness in AI** | 1 | AI decisions should be monitored and audited by humans, e.g., using FAIRness metrics (e.g., disparate impact), algorithms to mitigate bias (e.g., FAIR model training methods) and fair representation learning (e.g., adversarial debiasing).<sup>(9, 10, 17, 43, 50, 52, 57, 59, 60, 74, 78-80, 86, 87, 89, 90, 99, 101, 105, 106, 109, 111, 115, 118, 126, 127, 129, 131, 134, 139, 141, 143)</sup> | 33 |
|  | 2 | There are concerns about misconceptions and accountabilities around AI-driven decisions. These concerns can hinder AI implementation across organisations.<sup>(9, 10, 16, 28, 44, 46, 54, 57, 66, 74, 94, 99, 101, 103, 104, 107, 128, 130, 131, 133, 149)</sup> | 21 |
|  | 3 | Biased AI decisions can have serious consequences, e.g., prejudice towards or against certain social groups and erasure of diversity from data.<sup>(9, 10, 17, 43, 57, 60, 86, 90, 99, 101, 105-107, 109, 111, 127, 129, 131, 134, 141, 143)</sup> | 21 |
|  | 4 | There is a tendency to believe that automated decisions are bias-free, when in reality they are subject to the intrinsic biases of the humans involved in AI modelling.<sup>(9, 16, 17, 43, 60, 74, 85, 86, 90, 99, 101, 103, 105, 109, 111, 125, 131, 134, 143, 149)</sup> | 20 |
|  | 5 | Organisations should ensure they involve end-users and the public in AI decisions to obtain a balanced representation of views, expectations and concerns around the use and societal impact of AI.<sup>(9, 28, 51, 58, 60, 79, 102, 107, 117, 118, 130)</sup> | 11 |
| **Governance and ethical use of AI** | 1 | The EU Guidelines for Trustworthy AI recommend following a compliance criteria for evaluation of AI systems (transparency, privacy and data governance, technical robustness and safety, human agency and oversight, accountability, societal and environmental wellbeing, and diversity, non-discrimination, and fairness); the EU's AI4GOV project is developing a reference framework for an ethical and democratic AI, involving AI fairness monitoring, explaining AI decisions, training stakeholders and citizens, and boosting the regulatory compliance of AI models.<sup>(10, 26, 27, 68, 69, 98, 104, 118, 120, 122, 123, 128, 131)</sup> | 13 |
|  | 2 | Reports of ethical failures in AI use are increasing and enterprises face pressure to introduce sound and transparent controls for the systems they use.<sup>(9, 26, 38, 43, 54, 95, 103, 104, 109, 121, 122, 141)</sup> | 12 |
|  | 3 | GDPR now provides rules for information governance in AI, e.g., that human involvement is needed where AI is used to process and protect personal data.<sup>(4, 53, 64, 66, 99, 101-103, 107, 117, 118, 131)</sup> | 12 |
|  | 4 | Rapid advances in AI tools have outpaced the development of regulations and ethical guidelines, raising concerns about potential misuse and security of data.<sup>(10, 51, 90, 101, 107, 141, 152, 153)</sup> | 8 |
|  | 5 | The opacity of some algorithms ('black box AI') can have unintended social or legal consequences if not risk-reviewed (e.g., quality assured or regularly monitored) and subjected to human oversight.<sup>(28, 54, 61, 95, 107, 113, 122)</sup> | 7 |
|  | 6 | The IEEE Global Initiative on Ethics of Autonomous and Intelligent Systems for example have developed a set of ethical principles for AI, which are now widely recognised and used.<sup>(60, 78, 99, 122, 125)</sup> | 5 |

Extraction from the research funding or professional organisations was extracted in a separate Microsoft Excel spreadsheet containing: the RFO or professional body name; links to the relevant webpages; the date the information was accessed; and a summary of the relevant AI information (e.g., funding announcements, public statements, training activities, and guidance or initiatives).

## RESULTS

A total of 1,792 articles were retrieved from the two databases and 389 from grey literature searches (n=2,181). With 354 duplications removed and 818 assessed and screened for date relevance (2022–2024), 1,009 titles and abstracts, and then 216 full text articles, were assessed for eligibility (see **Fig 2**). Of the 216 full text articles, 108 met the eligibility criteria for inclusion and a further 14 were identified from manual searches, resulting in a total of 122 articles included in the scoping review (**see S2 Table: Full details of included studies**).

### Characteristics of the included articles

The characteristics of the included articles are provided in **Table 1**. All articles were reviewed against the classification framework(^6, 7^) and demonstrated inclusion of evidence on the 1) utility and potential of AI (n=112), or the 2) considerations and risks of AI (n=105). More than three quarters of the articles covered both areas of the framework (n=94).

All 122 articles were published between 2022 and 2024, with the majority published in 2023 (n=68). Almost half of the articles originated from Europe (n=54), with the remaining originating from the Americas (n=49), Asia (n=25), Australasia (n=7), the Middle East (n=4), and Africa (n=1). There were 85 peer-reviewed journal articles, with more than half reporting on original research (n=47), and a further 25 articles were perspectives, and 13 were reviews.

The literature on AI covered a wide range of sector and organisational settings (e.g., IT and software engineering, research and industry, healthcare, finance), and most of the reviewed literature was in the academic and big data/AI development space, with applications focused to data science, IT and software engineering (n=40) and/or research and industry (n=31). In 16 articles, the information covered was found to be relevant to multiple sectors or organisation types.

The evidence also covered a breadth of organisational AI application areas, pointing to numerous processes that could benefit from integrating AI systems. The most covered area was data management and security (n=91), followed by data analytics and visualisation (n=35), and decision-making and strategy (n=35). The least covered area was operations management and maintenance (n=5), which could be due to the evidence being more specialised to industry settings (e.g., manufacturing, production) and therefore having limited relevance to RFOs.

### Summarising the evidence

The evidence from the 122 included articles was summarised using the classification framework that was developed from existing AI/ML literature (see **Fig 1**).(^6, 7^) The summaries from the included articles were used to draw out key statements, which were then categorised into common themes and assessed for frequency (number of references contributing to each statement). **Table 2** presents the statements, frequencies and evidence sources regarding the utility and potential of AI, and **Table 3** presents the statements, frequencies and evidence sources regarding the consideration and risks of AI.

Interestingly, whilst mapping the evidence against the classification framework, an additional area around the importance of sharing efforts in AI, whereby articles focused on the value of collaborative working and partnerships between private sector companies (e.g., AI vendors), research institutions, and public administrations (e.g., including funding organisations) in the AI space. On this basis, key statements derived from article summaries promoting shared AI efforts across communities of interest were also extracted and presented in **Table 4** and **Fig 3**, along with the frequencies and evidence sources.

**Fig 3.** Mapping of the evidence across the utility and potential, considerations and risks, and shared AI efforts for research funding organisations (including themed areas under each)

**Table 4:** Summary of statements from the included articles on facilitating shared AI efforts in research.

| Key themes | No | Statements | No. refs |
| --- | --- | --- | --- |
| **Promoting inter-sector and inter-disciplinary collaboration in AI** | 1 | Effective transformations are needed to address aspects such as data cycles, skills, workflow efficiencies, available technologies, and necessary partnerships to ensure return on investment, growth, and sustainable social and economic development. Collaborations between cutting-edge data science research groups and the wider science communities is not yet common practice.<sup>(54, 56, 58, 64, 66, 67, 70, 74, 76, 80, 82, 86, 88, 89, 94, 103, 105, 106, 108, 109, 125, 139, 140, 144, 145, 147)</sup> | 26 |
|  | 2 | The most successful organisations will strike the right strategic balance based on careful calculation of risk, comparative advantage, and governance.<sup>(4, 9, 27, 36, 42, 44, 51, 54, 57, 64, 65, 69, 70, 86, 90, 91, 93, 94, 102, 110, 120, 121, 145, 149)</sup> | 24 |
|  | 3 | Organisations need to integrate diverse stakeholders into the development of AI systems to facilitate AI implementation, and to facilitate the co-development of AI systems.<sup>(27, 44, 49, 64, 69, 76-78, 84, 85, 90, 94, 98, 100, 101, 103, 105, 107, 121, 129-131, 145)</sup> | 23 |
|  | 4 | Interdisciplinary collaborative development needs to be inclusive at three levels: human values, humans' expression of intent and humans' abilities to create sufficient solutions that are sustainable across the research ecosystem.<sup>(9, 17, 28, 36, 38, 44, 51, 57, 63, 76, 78, 88, 108, 116, 120, 121, 132)</sup> | 18 |
|  | 5 | The AI field is currently dominated by private companies with profit incentives. There should be a public option designed to serve the public interest. There is constant pressure to improve efficiency, reduce costs, but the ability to use AI here is limited by costs of computational infrastructure and model development.<sup>(26-28, 39, 42-44, 46, 55, 56, 65, 75, 99, 100, 113, 125, 149)</sup> | 17 |
|  | 6 | Data information and sharing is critical for data management efficiency to improve societal demands, improve service delivery, and increase productivity (making it more dynamic and data driven).<sup>(26, 51, 53, 54, 64, 69, 74, 75, 82, 86, 101, 103, 105, 121, 123, 128)</sup> | 16 |
|  | 7 | Seek opportunities to join communities of practice; FAIR initiatives across Europe and the US, establishing the implementation, and adoption of FAIR principles in research; A joint statement on the AI Safety Summit (2023) from Academy of Medical Sciences for a collaborative, interdisciplinary approach to define how AI is governed, developed, deployed and used safely and ethically; US institutions (Stanford) supporting government efforts to build capacity to use AI through training initiatives; The Research Data Alliance guidance.<sup>(50, 59, 60, 74, 75, 78-80, 87, 99, 101, 105, 126, 127, 131, 133)</sup> | 16 |
|  | 8 | Aging data infrastructures and architectures directly impact AI/ML's sustainability performance. More than 75% of respondents stated that data architectures impact their sustainability performance.<sup>(50, 58, 61-63, 73, 76, 88, 120, 147)</sup> | 10 |
|  | 9 | AI implementation does not just happen. Organisations need a collaborative approach for the development of AI systems. i) integrate diverse expertise, ii) manage the translation of AI systems to business functions, iii) consider the workforce as an important stakeholder for AI implementation.<sup>(50, 53, 54, 57, 86, 94, 119, 141, 149)</sup> | 9 |
|  | 10 | More attention is required on public sectors role as an AI user and not just as a facilitator or regulator of AI technologies (facilitating and stimulating AI application within their own management and administrative practices).<sup>(27, 28, 42, 121, 125)</sup> | 5 |
| **Promoting leadership and support in data-driven practices** | 1 | Not all AI/ML technology are suitable or worth pursuing. The focus needs to be on what digital technology/ transformation will provide the most value to the organisation (and not test/pilot multiple tools). Research organisations need to maintain trust, and assess the risks and security concerns of AI implementation.<sup>(9, 10, 16, 26-28, 36, 42-44, 49, 52-54, 57, 60, 61, 64, 68, 69, 71, 74, 76, 88, 90, 91, 95, 98, 99, 101, 105-108, 116-118, 120, 122, 124, 127-131, 147)</sup> | 46 |
|  | 2 | Whilst AI is often discussed in replacing or automating human activities, human resources and skillsets are invaluable in AI capability.<sup>(1, 9, 17, 26, 42, 46, 51-54, 57, 64, 75, 79, 81, 84-86, 94, 95, 101-103, 108-123)</sup> | 40 |
|  | 3 | Gaining insight into how AI and digital technologies will impact research and science culture, including the use and application of AI-assisted tools to assist with research is starting to emerge (particularly around predictive analytics and natural language processing) but is still limited.<sup>(2, 17, 26, 28, 36, 42, 51-54, 60, 61, 64, 70, 72, 78, 80, 82, 84, 92, 94, 95, 105, 109, 110, 112, 121, 130, 145, 153)</sup> | 30 |
|  | 4 | Opportunities to stimulate the adoption of innovations, possibly when accompanied with financial incentives or other rewards to enhance management support and/or credible leadership with a vision for integrating new solutions in administrative processes.<sup>(16, 17, 27, 28, 36, 44, 50, 52, 54, 64, 65, 75, 77, 82, 86, 90, 93-95, 105, 110, 112, 116, 119, 139, 141, 145)</sup> | 27 |
|  | 5 | Explore ways to train staff on emerging technologies at all organisational levels and acquire human talent with expertise in AI, Big Data, and data science (crucial for emerging technological change).<sup>(9, 27, 43, 53, 54, 76, 77, 79, 83, 86, 94, 105, 109, 111, 114, 116, 119, 139, 147)</sup> | 19 |
|  | 6 | Data infrastructure needs to be well-designed, well-provisioned and well-staffed to enable data flow and allow for valuable insights to data quality enhancements. Gathering insights from several sources, institutions and organisations normalises and standardises data (enhancing data consistency and comparability).<sup>(26, 28, 42, 50, 52, 64, 76, 78-80, 82, 86, 90, 93, 105, 121, 132, 151)</sup> | 18 |
|  | 7 | When working with generative AI, context must be provided. Without context, AI is useless (e.g., receiving ChatGPT information that only dates back to 2021). If AI does not have the proper context to sift through data, then the data will be useless and the AI will be irrelevant.<sup>(9, 16, 39, 49, 53, 68, 75, 86, 92, 102, 104, 108, 117, 132, 142, 151, 153)</sup> | 17 |
|  | 8 | Data literacy, an ability to interpret and communicate with data, is crucial for effective data-driven decision-making in organisations. To enhance data literacy, organisations need to invest in training to educate staff about the value and usage of data. Investment in staff is required.<sup>(4, 27, 53, 54, 64, 75, 90, 92, 101, 111, 112, 119, 137, 141, 153)</sup> | 15 |
|  | 9 | All organisations and staff that develop, deploy, or operate AI should be accountable for the proper and accurate functioning; any AI operation should be transparent and explainable; AI technologies should be robust, secure, and safe throughout their entire lifecycle; AI technologies should be evaluated on a continuous regular basis.<sup>(42, 44, 61, 65, 66, 71, 95, 100, 103-105, 118, 119, 123)</sup> | 14 |
| **Promoting transparency and shared learning in AI** | 1 | AI governance, ethics and security requires a diverse group of stakeholders to provide a balance between digitalisation and risk management (e.g., the risks of AI tools and controls in place to manage them), to regulate developments and impacts of how AI tools are being used and modified to meet organisational requirements.<sup>(27, 44, 49, 69, 75-78, 84, 85, 90, 94, 98, 100, 101, 103, 105, 107, 109, 121, 129-131, 145)</sup> | 24 |
|  | 2 | Open source initiatives and frameworks to support open and shared learning, powered by large technology companies such as Google, Apple, Amazon (e.g., Tensorflow, Apache's Mahout, Spark).<sup>(2-4, 36, 39, 51, 53, 59, 60, 63, 64, 72, 79, 80, 83, 86, 93, 95, 99, 101, 104, 128, 147)</sup> | 23 |
|  | 3 | AI technology is still in the early experimental stage and there is a need for user validation, and investigative AI features are still at an early stage. More user validation and improvement with more annotated and testing data are required.<sup>(9, 52, 54, 60, 79, 97, 105-107, 109, 113, 114, 120, 121, 126, 147)</sup> | 16 |
|  | 4 | A shared infrastructure with openly accessible data within and across organisations will require a significant cultural change. Data literacy and data strategies are key components to achieving 'buy-in' from all levels of the organisation.<sup>(17, 36, 50, 52, 61, 64, 75, 82, 86, 100, 105, 112, 116)</sup> | 13 |
|  | 5 | Consider ways to manage unstructured data, place more emphasis on data infrastructure and architecture, assess ways to refine data using rigorous validation processes, regular reporting of effectiveness and areas for improvement, and consider multifaceted strategy as essential.<sup>(1, 42, 49, 52, 60, 62, 64, 70, 76, 79, 86, 112, 151)</sup> | 13 |
|  | 6 | Peer-reviewed publications should be linked to data, AI models, and scientific software to reproduce and validate data-driven scientific discovery. Openness of data and code is essential for the research community (e.g., researchers, RFOs) to test/pilot existing algorithms and the opportunities to build on these and leverage AI-ready (FAIR) data.<sup>(17, 36, 78-80, 109, 111, 116, 153)</sup> | 9 |
|  | 7 | Understanding the needs and gaps in data and AI models can promote sharing of knowledge and expertise at a rapid pace and scale. Unfortunately, such developments in focused areas such as Research and Development are currently lacking.<sup>(9, 27, 53, 54, 76, 79, 95, 101, 109)</sup> | 9 |
|  | 8 | Making use of open access publishing and platforms specifically for AI (e.g., F1000Research gateway) provides the opportunity for interdisciplinary publications and promotes research community alignment and a sharing research culture.<sup>(17, 36, 61, 78, 97, 98, 105, 116)</sup> | 8 |

The following sections provide a summary of the evidence captured from the included articles based on the review’s three research questions. The quality or assessment of the evidence was not explored as part of the review, and the key statements are not presented in order of priority.

### Utility and potential of AI for research funding organisations

Of the included articles, 112 described the utility and benefits (e.g., technological, functional, operational, and user focused) of implementing AI to support organisational, research or administrative processes. These articles either described the generic AI functions or systems (e.g., automation, generative AI, natural language processing) or pointed to specific tools (e.g., models, proprietary software or web platforms) that were either commercially distributed and ready to use (including open-source solutions) or remain in development (e.g., experimental ML algorithms).

It was clear from the evidence that understanding the utility and applicability of different AI technologies is complex, particularly due to the diversity of use across sectors and the different stages of digital transformation.(^49–54^) However, the evidence did provide several opportunities for RFOs to consider, particularly around the application, capability and implementation of AI. (see **Table 2**) The central focus was on AI/ML-powered tools and platforms, illustrating the transformative impact on data-driven organisational and/or administrative processes to enhance efficiency, innovation and decision-making.

### Advanced data management and analytics

The evidence offered several opportunities for using AI/ML platforms to enhance organisations’ abilities to manage and analyse large datasets, providing valuable insights for ways to improve customer relationships and operational efficiency.(^26, 49, 68, 71, 126^) AI-enhanced master data management and decision support systems also support data quality and can inform decision-making for stakeholders and facilitate long-term data management efficiency. For example, AI Ignite goes beyond building AI infrastructure, offering faster insights and operational efficiency, and empowering organisations to utilise the full potential of AI.(^72, 140^) While, AI-driven systems like Darktrace have also shown promising steps to enhance cybersecurity by preventing cyberattacks and are in turn driving the development of more robust public policies to govern AI cybersecurity.(^77^) Data mining algorithms and artificial neural networks are other AI solutions that can also inform organisations’ business intelligence models, and these work by enabling functions such as, data warehouse (e.g., Snowflake), data extraction (e.g., ETL) and report generation. (^55, 117, 136, 144^)

### Administrative task automation

Several ways of how AI can streamline public administration by automating services and processes, enhancing efficiency, and increasing public confidence was noted from the evidence.(^26, 28, 42, 54, 121^) For example, in accounting and law, AI has been used to automate repetitive tasks and enhance decision-making through predictive analytics and real-time data transparency, and therefore improving risk assessment and audit quality.(^1, 103, 133, 136, 141^) Conversational AI meanwhile showed to enhance efficiency and customer satisfaction by automating customer-staff communications.(^53, 65^) The evidence found that AI automation is predicted to save considerable time (40-50%) for administrative and analyst users, with this figure reportedly increasing to 80% as AI improves with more data. An Accenture analysis also determined that 40% of working hours across industries could be automated or augmented by AI adoption.(^86, 147^)

### Predictive and performance analysis

The evidence offered several ways on how AI-driven automation and predictive analytics can optimise resource-intensive activities and support sustainability by enabling remote work and real-time performance analysis.(^3, 52, 57, 147^) This is particularly relevant for RFOs who are keen to enhance project management, as AI has the potential to predict costs and risks, therefore harnessing productivity and complex data problems. In Human Resources (HR) practices, neural networks, NLP, and gamification tools could be leveraged to enhance recruitment, training, and performance, while ‘Empathetic AI’ can support leadership and HR processes through automation, improving employee support and organisational efficiency.(^57, 87, 111, 119, 124, 147, 149^) Other examples were shown in accounting, where predictive audit support enhanced risk assessment and audit quality, and in legal environments AI was shown to improve decision-making through real-time monitoring and data transparency.(^1, 92, 112, 120, 122^)

### Considerations and risks of AI for research funding organisations

Of the included articles, 105 described the considerations and risks (e.g., technological, operational, ethical and user focused) associated with ensuring organisational AI ‘readiness’ to assess capabilities and optimise organisation’s data infrastructure and management systems in preparation for AI/ML deployment (see **Table 3**). The evidence strongly indicated several organisational challenges that must be addressed for any business transformation or research activity involving AI/ML adoption to be credible. In particular, introducing or experimenting with AI technologies has raised concerns around data quality, issues with how data is shared, managed, and modelled for ML, and whether research and funding environments generally have the space, capacity, capability, and infrastructure to support and potentially mainstream the adoption of AI/ML for efficiency benefits.(^46, 50, 67, 116, 150^)

### Data storage and governance

From the evidence, several organisations were found to be implementing cloud and edge computing solutions to improve data architecture (e.g., how data is integrated, stored, and governed), manage AI/ML workloads, and cope with increasing data volumes and data types (e.g., text vs image, structured vs unstructured data). (^55, 64, 67, 70, 76, 100, 133, 145^) Cloud-based solutions like data lakehouse (^55^) and data mesh (^145^) can notably enable scalable and efficient data management, streamline data flows, and provide real-time insights for enhanced business performance. Examples of data warehousing solutions were also provided in the evidence (e.g., Hadoop, Microsoft Azure, and Apache Spark) and can reportedly support big data management and decision-making by offering comprehensive governance frameworks that enhance data quality, efficiency, and security. (^64, 73^) Cybersecurity was also highlighted in the evidence, particularly around how federal agencies and organisations are adopting secure cloud fabrics to ensure data governance, seamless connectivity, and secure access to data lakes. (^67, 151^) Other potential opportunities to enhance big data management and security included using distributed ledger technology like blockchain to securely manage digital assets (e.g., Syneos Health and startups like Granica) with comprehensive data management strategies for organisations adopting LLMs.(^98, 151^)

### Frameworks, models, and metrics

The evidence suggested that integrating standards, governance and ethics into workflows and AI models could alleviate the burden on innovation, and allow organisations to effectively utilise, regulate and govern AI.(^16, 51, 59, 79, 80, 117, 154^) A key consideration is having clear AI definitions to implement the necessary legal frameworks; developing fairness metrics for models; mitigating algorithmic bias; and implementing learning frameworks (e.g., IBM AI Fairness 360 (^134^)). Several frameworks and models were found in the evidence to regulate and verify the adoption and implementation of AI, such as the AI Ethics Orchestration and Automated Response (EOAR) framework (^99^), to ensure regular audits and testing of models; using platforms such as Open Scale to define, monitor, and tune the ethical behaviour of IBM AI models(^99^); the ‘Digital-Era Governance’ model for analysing the impact of digitalisation on administrations and(^125^); the International Science Council’s adaptive analytical framework (derived from the OECD AI Classification framework) which offers a comprehensive checklist for stakeholders to address AI impacts.(^69^) Other national-level efforts in this area included the UK Department for Science Innovation and Technology’s proposed regulatory framework for AI(^11^) to ensure responsible innovation and research-driven initiatives in the US and Europe (e.g., HPC-FAIR, AI4GOV project) aimed at redefining and applying FAIR principles to maximise the impact of data investments and monitoring AI for bias or non-compliance. (^78, 131^)

### Enhancing big data analytics

Several cloud service providers were suggested such as AWS, Azure, and Google Cloud Platform (GCP) to enhance data management and AI-powered analytics in the financial sector, offering scalable, flexible solutions for handling large data volumes, enabling real-time decision- making, and improving operational efficiency. (^70^) In addition, edge computing can pinpoint and locally process the data necessary for analyses, providing scalability, real-time data analysis, interoperability and confidentially for emerging ‘Internet of Things (IoT)’ (e.g., sensor and ICT-based) solutions.(^71^) Developments in Blockchain technology were also shown to assist AI algorithms in managing big data in healthcare, and can help monitor data and assess the integrity of analytical outputs. (^97^)

### Data standards and research tools

The evidence reported on the development and use of data standards for AI-driven research, as well as data-centric initiatives to promote AI data quality, facilitate big data management in research, and ensure that data is more findable, accessible, interoperable and reusable (FAIR) across organisations, institutions and platforms. (^50, 59, 60, 78–80, 154^) For example, the FAIR4HEP collaboration promotes FAIR data principles for the reuse of experimental data within and between institutions(^80^); the Research Description Framework (RDF) provides semantic web standards for consistently describing, integrating and exchanging metadata (^59^); and a proposed computational framework for quantifying the FAIRness of AI models and adopting AI-ready datasets has been developed to enable AI-driven scientific discovery.(^79^) From a business perspective, knowledge graphs and semantic web standards were reported to enhance AI implementations by automating workflows and facilitating complex queries(^132^) and semantic web standards also ensure data compliance with FAIR principles, enabling efficient data governance and retrieval.(^59^)

### Technical infrastructure

The evidence provided important considerations for organisations who wish to be **‘**AI ready’ in their capabilities(^90, 94, 109, 127, 129, 138^), such as upgrading outdated infrastructure, maintenance and management systems(^3, 65^); harmonising business processes; standardising and validating master data; and enhancing IT agility (e.g., by adopting Cloud) and ensuring, competitiveness, and resilience.(^65^) However, it was also evident that upgrading maintenance management systems requires adopting big data, cloud computing, and automation, managing vast data efficiently, addressing sustainability issues, and ensuring robust cybersecurity.(^3^) Investment in data availability, quality and careful evaluation of cloud resources for model training are also crucial (e.g., for implementing generative AI models) and adopting cloud-based solutions in particular (e.g., Cloudera, Azure cognitive services) is advantageous as these support routine tasks; expedite report preparation; enable quicker strategic decisions; and promote data democratisation.(^65^)

### Guidance on ethical AI

One of the key considerations from the evidence was for the need for countries and organisations to adopt ethical guidelines around AI tools, and LLMs like ChatGPT in research. (^66, 107, 113, 117, 128, 131, 153^) Several factors were mentioned around AI’s capacity to manage and analyse large data volumes (also enhancing data integrity), the need for human oversight to mitigate risks (e.g., reputational, legal, and regulatory)(^113^); ensuring responsible use of AI in research funding and management(^36^); and ensuring the protection of democracies and the risks to human equality posed by big data models (e.g., bias against certain groups).(^131, 134^) Regulation and accountability in big data is essential, and there is a need for public sector organisations to consider the historical barriers to innovation; embrace technological changes; leverage data to meet public needs; and replace outdated systems.(^28^) However, any integrated AI solutions must uphold existing rules regarding data integrity, ensure proportional and transparent data collection, and maintain confidentiality of sensitive data (e.g., patient, public, and research data).(^4, 51^) Several regulatory efforts were reported, focusing on national AI model risk management frameworks, enhancing governance, and ensuring model lifecycle updates.(^107^) For example, The EU guidelines on ethics in AI, emphasises the need to ensure AI systems meet criteria for transparency, privacy, and fairness(^4^); the United Nations Educational, Scientific and Cultural Organization’s (UNESCO) recommend establishing a universal framework of ethical AI principles and actions(^66^); the EU’s AI4GOV project is developing a reference framework for ethical and democratic AI(^131^); and the AIDE Project (Artificial Intelligence in Healthcare for All), a UK and Japan collaboration to identify and address public concerns regarding AI use in healthcare.(^130^)

### Facilitating shared AI efforts

In addition to the above, the evidence also strongly indicated that facilitation of shared AI efforts through collaboration and partnerships is important in achieving AI readiness and successful implementation in the research sector. Fostering transparency and shared learning between the key stakeholders in research (e.g., researchers; RFOs; academic institutions; public and private organisations; public representatives) is crucial for a) understanding the real-world use for AI/ML in administrative research and research management practices, and b) enabling development of and access to solutions that can be transferable across several organisational and data management contexts.(^10, 26, 28, 31, 33, 42, 121^) The statements below (see **Table 4**) provide a summary around the importance of an open culture and shared AI efforts between organisations and sectors who have a role to play in research.

### Developing organisational AI strategy

From the evidence it was clear how understanding data value and trends is crucial for developing AI capability. Although understanding data requirements are closely tied to government policy (and initiatives), adherence to ethical standards, and availability of financial, knowledge and technical resources, there is a need for organisations to carefully consider AI solutions and develop data strategy plans that encompasses both AI and data management practices.(^53, 92^) Investing in data strategies, with the significance of data literacy requirements, a key consideration for organisations is the potential to invest in collaborations and partnerships to facilitate AI implementation.(^26, 27, 53, 54, 64, 66, 67, 70, 74, 86, 89, 94, 109, 119, 125, 139, 145, 147, 149^) Digitalisation strategy plans need to identify key performance indicators and effective adoption of ethical AI business practices, which in turn should involve having the right organisational structure; an interdisciplinary working culture; communication; management support; training; appropriate measurement and reporting mechanisms; and enforcement.(^9, 27, 43, 54, 76, 79, 83, 86, 94, 109, 111, 114, 116, 139, 147^) The evidence also suggested that AI application management and AI-driven platforms need to facilitate stakeholder interactions and continuously innovate in rapidly changing environments.(^44, 77, 98, 107^) LLMs in particular require a strategic roadmap for effective integration, with careful assessment of applications, techniques, and regulatory considerations. In financial management, the strategic roadmap should involve AI being implemented in stages and only follow once data management infrastructures have been upgraded.(^50, 67, 90^)

### Monitoring AI transparency

Strengthening accountability in AI (‘explainable AI’) requires human oversight and involvement throughout the model lifecycle, as well as engagement with internal stakeholders and AI users to avoid automation bias.(^101^) The development and adoption of explainable AI was mentioned across several organisational settings, with particular focus on the transparency and interpretability of AI technology.(^61, 86, 95, 101, 103–105^) For companies developing powerful AI systems (e.g., OpenAI), this means taking appropriate steps to involve independent members to ensure democratic AI governance and to prevent misuse.(^104^) Organisations, particularly in healthcare or research settings, need to ensure well-documented datasets for all AI technologies; identify user groups at risk of disparate performance or harm; justify appropriate dataset use; evaluate performance across groups; report limitations or misuse; and develop risk mitigation plans throughout the AI technology lifecycle.(^26, 42, 52, 64, 87, 104, 107, 151^) The evidence also highlighted how model training and lifecycle development (e.g., for LLMs) need to explore incorporating FAIR data principles at every stage to generate FAIR-compliant datasets.(^50,^ 60, 74, 75, 78–80, 99, 101, 105, 126, 131, 133, 155)

### Increasing AI partnerships

Several initiatives and alliances were reported in the evidence, particularly around forming partnerships to ensure return on investment and sustainable social and economic growth.(^64, 74, 86, 94, 103, 125, 139, 154^) Public and private sector collaborations can offer comprehensive support solutions across various stages of AI implementation and co-develop AI systems that are intrinsically inclusive of human values and humans’ ability to create sufficient solutions that serve the public interest.(^9, 17, 26, 28, 42, 44, 51, 120, 121, 132^) Initiatives such as the Responsible AI UK (https://www.rai.ac.uk/) and the European Commission’s joint Research Centre’s Artificial Intelligence Watch (https://ai-watch.ec.europa.eu/index_en) were two examples seen in the evidence to enhance potential collaborative partnerships as well as strengthen data management and administrative practices in research.

### Developing AI knowledge and skills

A key highlight from the evidence focused on staff training; staff development; career growth; and building talent that is adaptable to the rapidly changing technological advancements across and within different sectors and organisational settings.(^50, 57, 86, 149^) The evidence suggested how organisations need to invest time and resources into addressing the skills gap in order to successfully implement AI and understand the level of support required for the adoption of AI.(^50, 53, 119^) Although there was some evidence around the implementation and adoption of AI to streamline processes and reduce bias, data literacy and data strategies are key components to gain ‘buy in’ from all levels and sections of an organisation.(^17, 64, 75, 112, 116, 119^) Upskilling and offering AI education will be required across whole organisations, especially as AI-driven scheduling software and intelligent assistants are increasingly being implemented to reduce management and administrative burdens.(^9, 37, 94, 134, 141^) In educational settings, promoting knowledge around AI and adequate resources is required to support AI management and learning, facilitate staff willingness to adopt new digital skills and to reduce instances of unethical AI use that could have implications on academic integrity.(^20, 153^) In more business focused settings, data leaders are already capitalising on AI, or expecting increased interest and investment in AI technologies, by upskilling their employees in AI/ML, and focusing more on improving data literacy and data readiness for AI in future data strategy plans.(^92^)

### Evidence from funding and professional organisations in research

To gain further insight into the priorities and positions of key funding and professional organisations with respect to AI, we reviewed 13 webpages of research organisations (**See S3 Table: Key research funding and professional organisations on AI: announcements and activities**). It was clear that several organisations are initiating funding opportunities that involve AI, and this was particularly evident from organisations focused on healthcare. For example, the UK Research and Innovation’s (UKRI) funding opportunities to accelerate new treatments and therapies using AI technologies, and the NIHR and Wellcome funding schemes to support healthcare research involving big data (https://www.nihr.ac.uk/explore-nihr/funding-programmes/ai-award.htm). Several key areas were noted from the review of a purposive sample of relevant organisations, which are reported below.

### Ethical conduct

High on RFO’s agendas is ensuring responsible use and addressing the ethical challenges that AI presents, which had led some organisations such as the Canadian Institute of Health Research (CIHR) and UKRI councils to set up leadership groups and advisory committees in ethical AI/ML (e.g., Engineering and Physical Sciences Research Council (EPSRC), and the AHRC and Economic and Social Research Council (ESRC) have partnered to create a leadership team to drive the UK’s responsible and trustworthy (R&T) AI agenda). Other RFOs, like UKRI’s Arts and Humanities Research Council (AHRC) and the Academy of Medical Sciences (AMS), are exploring ways of including ethics in the research and development of AI technology, recognising the need for strengthening collaborations and partnerships with other organisations and technical experts to ensure that AI is used safely, effectively and ethically.

### Security and confidentiality

Another area high on RFOs agenda focuses on security and confidentiality of personal data when using AI tools for peer review or grant applications. As a result, a joint statement from the UK’s Research Funders Policy Group, which include Wellcome and the NIHR, issued caution against the application of LLMs such as ChatGPT. The National Institutes of Health (NIH) have gone further by prohibiting peer reviewers from using any AI technologies for analysing or formulating their reports on grant applications and contract proposals (e.g., NLPs, LLMs and other generative AI tools). In a response to these concerns, several organisations such as the UKRI’s Challenge Fund are supporting service industries to use AI and data analytics in the security of their computer infrastructure.

### Equitable AI

Preventing and mitigating biases were mentioned by several RFOs like the CIHR, who highlighted in a report from the *AI for Public health Equity Workshop* in Toronto planned efforts to promote equitable AI and equip public health sectors in Canada with new methods to advance health equality as a priority area (Public Health: IPPH Strategic Plan 2022-2026). The Gates Foundation are also focusing on access and equity in AI work and have established a task force to ensure AI usage is safe, ethical and equitable worldwide. Furthermore, the NIHR have announced their Artificial Intelligence and Racial and Ethnic Inequalities in Health and Social Care funding call to conduct work ensuring that AI research meets the needs of ethnic minorities (https://www.nihr.ac.uk/explore-nihr/funding-programmes/ai-award.htm).

### AI knowledge, skills and resources

Although, several RFOs highlighted how they are assessing the need for staff to enhance their research skills through interrogation of AI, they are addressing this in different ways. For example, the NIHR ARC North Thames Academy have set up an AI in Systematic Reviews Masterclass for healthcare and government staff (https://www.arc-nt.nihr.ac.uk/news-and-events/2024/feb-24/the-use-of-artificial-intelligence-ai-in-systematic-reviews-masterclass/), while the NIHR CRN have developed an AI e-learning course in partnership with Imperial College London (https://www.nihr.ac.uk/news/artificial-intelligence-e-learning-launched-for-researchers/31387).

### Big data in research

The Association of Research Managers and Administrators (ARMA) first national survey of research offices predict that research management practices are poised for increased use of AI and big data in the near future. Similarly, the Cancer Research UK’s (CRUK) Grand Challenges Fund demonstrates that researchers are also looking to harness big data and AI for medical data analytics for early detection and treatment of cancers. On a larger scale, the EU has announced a Digital Strategy to establish standards in data and invest in AI, while major funders such as the NIH are establishing data science and AI programmes to design and develop intelligent data-driven AI-based medical screening tools.

### Public scrutiny

Several organisations have examined the use of and implementation of AI to ensure that the public’s best interest are protected when using AI, while also considering (where applicable) regulations such as the EU Act (e.g., the European Commission AI Watch monitor the development, uptake and impact of AI across Europe through a regulatory framework based on human rights and fundamental values). Outside of the EU, the Gates Foundation are in efforts to make AI technology broadly accessible to all to transform how people communicate, work and learn, and the Joseph Rowntree Foundation are coordinating discussions around harnessing AI for public good. Other examples of promoting public scrutiny in AI include the Association of Medical Research Charities (AMRC) and their annual Benevolent AI award to help accelerate new treatment and therapies using AI, the CIHR have two initiatives through their Transforming Public Health IPPH Strategic Plan and AI Public Health Equity Workshop, and the NIHR’s plans to apply real-time operational modelling to maximise care pathways (https://www.uclhospitals.brc.nihr.ac.uk/criu/research-impact/artificial-intelligence-and-machine-learning).

## DISCUSSION

With this review, we addressed the complexities surrounding AI-powered digital transformation, drawing on evidence from a range of sectors, user contexts and AI application domains to highlight current and emerging applications of AI, their potential benefits, and the key considerations and risks for RFOs. Much of the evidence was solution-focused, using either tools, frameworks, models or general recommendations on how to prepare for and implement AI across diverse organisational contexts. A good proportion of the literature was from original research or research perspectives published in peer reviewed academic journals. By using a classification framework to capture the utility and potential, and considerations and risks of AI(^6, 7^), we identified key aspects for RFOs who are already in the process of, or are looking towards, adopting AI/ML within their organisation (or for their research or funding activities). In doing so, we also identified a compelling body of evidence indicating the need for more shared efforts in AI, and the importance of an open and collaborative culture between research stakeholders and communities of interest to improve AI capabilities, knowledge, and organisational strategies.(^26, 28, 42, 50–54, 60, 64, 78, 94, 95, 101, 121, 143^) It was clear from the evidence that organisations are leveraging AI to build strategic capabilities, seek opportunities to remain competitive, and find ways to enhance consumer experiences. For sectors like research, this area of digital transformation has been comparatively slow, due to the hesitancy around ethics and regulation, and the uncertainty around AI’s utility and the capabilities required for AI implementation. However, the level of hesitancy from RFOs is largely the result of a lack of clarity around AI-driven transformation, concerns around the ethical and societal implications of AI, and the need for more empirical evidence to develop governance frameworks, models and guidance to enable the responsible use of AI for data management in the public sector.(^54^)

### Current and emerging use of AI

Adopting and successfully implementing AI is complex, particularly when comparisons are made between the private sector (the ‘facilitator’ of AI) and the public sector (seen as the ‘regulator’).(^26, 28, 42, 121^) Added to the complexity is the need for organisations to assess the relative merit and effort required to implement AI technologies, which are particularly important in considering the readiness and capabilities of AI, and what is needed to address the unique challenges of AI adoption.(^9, 28, 51–53, 64, 92, 94, 121^) A large proportion of the evidence was relevant to the research sector, particularly around improving the quality of research data management (making it FAIR and verified for AI/ML training), offering important considerations for RFOs who collect research data to monitor and report on the impacts of funded research (for accountability and to secure further investments). RFOs manage high volumes of data both directly from awards and from other sources, and they engage in analytics to manage, monitor and evaluate their organisations funded portfolio. The evidence has shown the potential benefits and capability of AI, and how it could enhance efficiency and insights in these processes, particularly with ML. Standardising research data management and improving the quality of research data is needed to ensure accuracy of ML models applied to business and research processes, and ultimately the success of outcomes. However, it is also important for research in the wider context, based on reports of increasing research bureaucracy caused by lack of alignment in standards and data systems in digital research environments.(12, 13, 50, 76, 78–80, 132, 151, 154, 156)

The focus of the available evidence on AI in research for academic use may be explained by the fact that this is where AI is mostly used due to free access tools such as ChatGPT(^6, 18, 20, 153, 157^), and subsequently raises several ethical concerns (e.g., data storage, data privacy and security, and the use of information). Although several efforts were noted from the literature(^26, 101, 107^) (e.g., ‘Explainable AI’ techniques such as LIME and SHAP) these may not necessarily anticipate all ethical and governance issues that may come up when AI is used more widely across public administrations (where accountability and public perception is more crucial).(^9, 26, 27, 43, 87, 90, 101, 111, 154^) RFOs require high quality evidence to inform their own practices with regards to the adoption, implementation and capability of AI use, particularly when some funding organisations are taking a position on the use of AI during the funding application process (e.g., peer review and grant writing).(10–12, 24, 32, 36, 79, 158)

### Potential benefits of AI for RFOs

To gain a comprehensive picture of the AI landscape, we considered existing AI applications, new developments in technologies and emerging uses for AI that could be applied to organisational contexts in research (e.g., research data management and administration). There was emphasis on exploring how AI could be leveraged to reduce research bureaucracy, particularly administrative burden for RFOs, based on widespread reports of increasing pressures in research to demonstrate impact and value of investment.(^31, 32^) However, the increasing effort also extends to researchers, where most data management processes are manual and time-consuming.(^98^) AI implementation could have the potential to reduce prompt-like queries, and develop Edge AI applications and NLP models for human-AI interaction to shift towards enhanced insight and reduction in burden for researchers.(^53^) Despite the advantages of AI applications though, they are not a replacement for human intelligence or critical thinking; they are tools to encourage more streamlined approaches to data management, research and educational processes.(^53, 120^)

It is also particularly challenging for RFOs, given the rapid speed of AI/ML development, to address issues around performance, fairness, bias (e.g., ‘AI hallucinations’ which is when AI models generate incorrect, misleading or provide false information) and maturity of AI technologies. There are increasing efforts focusing on the integration and capability of AI, such as the Responsible Artificial Intelligence UK (https://rai.ac.uk/) and the European Commission’s Joint Research Centre’s Artificial Intelligence Watch (https://ai-watch.ec.europa.eu/index_en), which could facilitate and foster greater reliability for the adoption and implementation of AI in research data management.(^27, 28, 42, 43, 106, 116, 121, 154^) Solutions are still needed to not only manage the current demand placed on RFOs, but also where AI/ML tools could serve organisations and the wider research community (e.g., by reducing duplication and facilitating monitoring, reporting and evaluations) (**see Box 1** for a summary of the evidence on the key considerations for optimising AI implementation for RFOs).

#### Box 1.

##### Future considerations for AI implementation for research funding organisations

- Harnessing AI for efficiency and innovation in research, and ensuring its responsible use across research contexts, requires a multifaceted approach and commitment from all relevant sectors and organisations, including the AI/ML experts and industries (e.g., Amazon, Google, Meta, Microsoft and OpenAI) that develop AI tools.
- Industry 4.0 has revolutionized organisational processes and capabilities, with big data and advancements in AI (ML, LLM, NLP, IoT) now offering practical business solutions and insights to assess the need to improve AI capability within organisations (not just focusing on AI capability, but also the consideration of the fragility of the AI capability).
- Edge AI applications offer a solution for AI explorers, looking to develop and explore possibilities and siloed structures, allowing for data privacy and security and mitigate against concerns on data accessibility.
- The growth and speed at which AI has advanced over the last few years demonstrates the need for embedding openness, transparency, and curation for digital transformation.
- AI technology can have the potential to reduce research data management and administrative burden, enhance productivity and free up time to dedicate and redirect focus and attention to value-added activities that require complex decision-making.
- With the adoption of AI, it is important to consider the productivity of AI but also transformational AI (e.g., the creativity and competitive advantage by completely reforming processes and enabling new systems such as moving to the cloud to enhance integration and interoperability).
- Lack of understanding of the value, impact, and sustainability of AI adoption remains a significant gap in the evidence. There is a need for evaluation studies to understand whether AI technologies/ AI-enabled innovations can achieve the intended goals, and help to build trust in the use, applicability, and adoption for the application of AI for RFOs.

### Considerations and risks of AI for RFOs

Concerns over current data quality, data management practices, and the implications for AI/ML model accuracy and effectiveness, are common across sectors, organisations and disciplines. (^78, 134^) The overall risk with using systems that are inaccurate, biased, or compromise human rights (e.g., privacy, accountability, security) is that users and leaders will lose trust in AI and not look to harness potentially useful technologies that could reduce the burden on innovation.(^123^) Although, the negative outcome can be avoided by ensuring human involvement throughout the AI model lifecycle, it is imperative for stakeholders and end-users to be actively involved in decision making throughout the initiation, and development of establishing systems for monitoring and evaluation purposes.(^92^)(see **Box 2** for a summary of the evidence on the key considerations to develop trust in AI implementation) There are also requirements for regulatory efforts to ensure safe and responsible AI innovation, particularly due to ethical and governance concerns around data integrity, data collection and confidentiality. Data initiatives and insight are constantly shifting (and at a rapid pace) and there is a clear drive for organisational and government strategies to engage in the long-term sustainability of AI technologies.(^4, 91, 93, 146^) These investments in AI are predicted to be long-term and sustainability should not be overlooked by focusing on cost alone. (^150^) Moreover, integrating AI technologies requires new initiatives to modernise existing research data management practices; whilst that may come at a cost, it will inevitably produce greater efficiencies and optimise reduction in operational costs resulting in longer term efficiency gains for RFOs.

#### Box 2.

##### Future considerations for developing trust in AI implementation

- Ensuring best practice by following the latest developments and evidence in the ethical and responsible use of AI.
- Working with other funding organisations and government departments to further develop guidelines in AI Ethics and Governance, determining where they should be tailored to organisations or sectors and where establishing universal standards is needed and feasible.
- Developing in-house protocols and risk assessments for assessing where automation and AI can best support human tasks, increase efficiency, or enhance decision-making.
- Developing in-house monitoring systems (and metrics) to ensure transparency and fairness of AI models and to avoid systemic biases.
- Develop strategies to ensure compliance of research data with FAIR principles in line with other funding organisations such as Cancer Research UK and the National Institutes of Health.
- Build on existing capacity and capabilities to enable AI data preparation, model testing and implementation, where needed bringing in experts in data science and data management and ML/AI.

### Shared AI efforts across sectors

There was a particular emphasis on the need to promote openness and transparency through communities of interest (e.g., different sectors, settings and organisations), particularly when there is a need to reduce the burden on innovation, and facilitate trust with AI technologies.(^16, 81, 83^) Facilitating and fostering transdisciplinary collaborative opportunities across diverse cultural domains could offer greater reliability of potential applications and promote a more collaborative, data-driven approach to further advance the adoption, capability and implementation of AI/ML technologies (from a collective viewpoint rather than from one organisation) (**see Box 3** for a summary of the evidence on the key considerations around shared AI efforts). According to the International Science Council, there is an ontological gap between AI principles and their integration into regulatory and governance frameworks, requiring a systems approach that includes the scientific community and considers the broader implications of AI for science and society.(^69, 78^) Effective regulation will necessitate risk-informed decision-making across multiple layers and stakeholders, including RFOs. The adaptive analytical framework could serve as an example for RFOs as they assess multiple layers of impact and are looking to develop a more systematic approach to monitoring and reporting outputs, outcomes and impacts.(^69, 78^)

#### Box 3.

##### Future considerations for shared AI efforts

- Develop partnerships to be able to explore the strategic and organisational challenges around the use and implementation of AI for research funding organisations, paying particular attention to establishing greater community participation with a long-term investment.
- The benefits of AI and ML will remain limited to research funding organisations and institutions unless more research explores the interpretability of the effectiveness of AI (including the implications for several stakeholders such as staff, researchers, end users and the public).
- AI and data literacy requirements and training are required to understand the broader context and content about the application of AI (AI literacy framework).
- Develop and seek opportunities to experiment and pilot where AI can be applied to specific processes and systems in research management practices (including regular evaluation and interrogation on these technologies).
- Seek to establish a framework for responsible use of AI/ML in research management practice to enable funding organisations to be aligned and have a unified approach in business process (understanding that there will be variations across funding organisations). Promote openness and transparency through communities of interest and seek to share practices across funding organisations to enable greater transparency and generalisability.

With much of the literature in the data science space, RFOs may need to assess and consider how to upskill their workforce and seek collaborations with software companies and data science experts/ML researchers.(^9, 27, 54, 76, 83, 86, 94, 105, 109, 111, 139, 147^) Organisations could also engage with data scientists for AI expertise and to post-process their datasets, (clean, assemble and validate the data). Where AI is experimentally used on imperfect data, this reduces some processing effort and guides future data collection practices. These shared decision-making and learning practices across sectors could ultimately help foster and drive business performance and transformation, and by implementing strategies to enhance data management would be a competitive advantage in today’s data-driven world.(^26, 28, 42, 79, 80, 90,^ 93, 105, 121, 132, 151)

### Study strengths and limitations

The main strength of the review was the extensive coverage of sectors across varied organisational settings and including a review of 13 research funding and professional organisations. Using a framework to map the evidence provided a clear interpretation of where and how AI has been and is being applied and adopted across a range of settings to determine the potential benefits and opportunities, and consideration and risks associated to implementing AI technology for RFOs. However, a limitation to the review was that scoping reviews are restricted to only mapping the evidence and do not assess the quality or risk of bias of the literature. Although we mapped the evidence by research methodology (e.g., original research, perspective, review) due to the wide scope of the review (not limited to a particular sector, discipline or field) it was clear how some research methodologies were not as developed or consistently defined as one would expect in healthcare or food and environmental settings. The review only covered a three-year period due to the rapid pace of evolving evidence (also noting that literature published prior to 2022 was more conceptual and focused on algorithm development only), which could also be seen as a limitation to the review. Although, there was some international coverage from the included articles, the majority were from Europe and Americas, which could have introduced regional biases. However, it could also be that there was a lack of evidence from these regions, given the coverage of the databases and grey literature searches.

## Conclusions

Big data and AI has been a subject of intense public debate, but when it comes to applications and considerations for RFOs there was a clear evidence gap in the literature. This review has provided a ‘snapshot’ of the current evidence, across diverse sectors and settings, at a time when RFOs have been undergoing digital transformation and reviewing efforts to address research bureaucracy and barriers to collaborative and unified research management practices.(^31, 32^) Whilst previous research has focused on the interoperability in collection of research data, use of platforms, compliance and reporting requirements, there are still areas of concern to reduce current research burden (inclusive of researchers, institutions, funding organisations), which could also unintentionally introduce or increase administrative efforts.(^33^) However, in order for RFOs to be good custodians of public money, there will always be some form of administrative effort required around monitoring and reporting, in order to demonstrate the tangible value and benefit of research (from both RFOs and researchers).(^26, 28, 42, 54, 121^) Adopting and employing technological advances such as AI/ML could be a solution to change the research funding landscape. However, due to the nature of RFOs, and the increasing emphasis on accountability, societal impact, research integrity and transparency, there is a need to consider the published evidence around AI, with specific emphasis on its application, potential, and associated risks in the context of funding activities. With the increasing rise of digital transformation, advanced analytics and data-driven decision-making means a concerted research effort into understanding the opportunities, capabilities and risks of AI is needed more now than ever. Collaborative efforts are required to ensure such technologies are not only accessible and ethical but also support adequate advancements within organisations across different sectors to prepare, educate and train both staff, wider customers and users of AI in the long term.

## Supporting information

S1 Appendix search terms and keywords

S1 Table search strategies

S2 Appendix PRISMA checklist

S2 Table full details of included studies

S3 Appendix abbreviations

S3 Table key organisations

## Data Availability

All relevant data are within the manuscript and its Supporting Information files. They are also available from the OSF https://osf.io/d8bfn/

## Acknowledgements

We would like to thank Nancy Beckett-Jones from the University of Southampton Library services in developing the search strategies for the review, Sophie Hall for conducting Overton searches for funder literature on AI, and Gi-Yian Yiu for identifying and collating AI/ML tools described in the literature.

## Supporting information

**S1 Appendix.** Search terms and keywords

**S2 Appendix.** PRISMA checklist

**S3 Appendix.** Full abbreviations

**S1 Table.** Search strategies

**S2 Table.** Full details of included studies

**S3 Table.** Key research funding and professional organisations

## Registration

This study is registered on the RoR Registry and Hub (study number 3292) and can be accessed here: https://ror-hub.org/study/3292/

## Notes

### Competing Interest Statement

The authors have declared no competing interest.

### Funding Statement

This research was internally funded by the National Institute for Health and Care Research (NIHR) Coordinating Centre, based at the University of Southampton. The views and opinions expressed in the discussion are those of the authors and do not necessarily reflect those of the NIHR or the Department of Health and Social Care. The NIHR had no role in the design, collection or conduct of this study, nor the decision to publish or the preparation of the manuscript.

