## Supplementary material for "Exploring the potential benefits and challenges of artificial intelligence for research funding organisations: a scoping review": S1 Appendix search terms and keywords

S1 Appendix: Search terms and keywords used

| Search topics/area | Search terms |
| --- | --- |
| Artificial intelligence | Artificial intelligence  Generative artificial intelligence  GenAI |
| Management and administration | Data management  Management information system  Administration and organisation/organization  Administration and planning |
