## Supplementary material for "Exploring the potential benefits and challenges of artificial intelligence for research funding organisations: a scoping review": S1 Table search strategies

| **Web of Science (01/02/2024)** | **Results** | **ProQuest (01/02/2024)** | **Results** |
| --- | --- | --- | --- |
| (TI=("artificial intelligence" OR GAI OR AI OR GenAI )) OR AB=("artificial intelligence" OR GAI OR AI OR GenAI)  (TI=("data management" OR "management information system*" OR administration and organisation OR administration and organization OR administration and planning)) OR AB=("data management" OR "management information system*" OR administration and organisation OR administration and organization OR administration and planning)  #1 AND #2  #1 AND #2 and 2024 or 2023 or 2022 or 2021 or 2020 or 2019 (Publication Years) | 196,625  69,298  805  **646** | abstract("artificial intelligence" OR GAI OR AI OR GenAI) OR title("artificial intelligence" OR GAI OR AI OR GenAI)  title("data management" OR "management information system*" OR "administration and organisation" OR "administration and organization" OR "administration and planning") OR abstract("data management" OR "management information system*" OR "administration and organisation" OR "administration and organization" OR "administration and planning")  [S1] AND [S2]  [S1] AND [S2]Limits applied | 641,357  72,509  1,478  **1,146** |
