## Supplementary material for "Exploring the potential benefits and challenges of artificial intelligence for research funding organisations: a scoping review": S2 Table full details of included studies

| Record metadata | | | Summary of relevant AI information | | | | Classification framework checklist | |
| --- | --- | --- | --- | --- | --- | --- | --- | --- |
| **Author and Year** | **Source type** | **Country/ies** | **Organisational setting/s or sector/s** | **Organisational area/s of AI application** | **Summary of the issues** | **Summary of the solutions** | **Evidence on utility and potential of AI (Y/N)** | **Evidence on considerations and risks of AI (Y/N)** |
| Anonymous, 2022a | Magazine | USA | Data science, IT and software engineering (Dell Technologies, Cloudera) | Data management and security; Data analytics and visualisation | Organisations face 30-40% annual data growth and consequent data protection and management burden, which often leaves valuable “dark” data unanalysed. | AI and ML-powered data analytics platforms enable organisations to gain insights at scale, enhancing customer relationships, operational efficiency, and innovation. Public clouds (e.g., Cloudera Data Platform) and on-premises data centres offer speed and cost efficiency. | Y (Technological advances, Operational efficiencies, User focus, Functionality) | Y (Technological) |
| Komprise, 2023 | Report | UK and USA | Business, enterprise and finance; Data science, IT and software engineering (IT and business companies) | Data management and security | IT and business leaders generally permit employees to use generative AI, but 66% are concerned about data governance risks and AI vendor transparency, especially with respect to risks of unstructured data | Strategies to mitigate AI risks includes understanding data value and trends and options for data storage and monitoring capacity issues (e.g., Amazon Web Services) | Y (Technological advances, Operational efficiencies, User focus, Functionality) | Y (User, Operational, Technological, Ethical) |
| Anonymous, 2023 | Magazine | USA | Public administration, policy and legislature | Data management and security | Cloud technology has many benefits but also poses security risks, especially when it comes to protection of sensitive data | Federal agencies are adopting secure cloud fabric that ensures data governance, seamless connectivity, secure access to data lakes (e.g., for analytics) and supports AI infrastructure to enhance customer experiences and optimise workloads | Y (Technological advances, Operational efficiencies, Functionality) | Y (Technological) |
| Flexera 2023 | Report | USA, UK and Europe | Business, enterprise and finance; Data science, IT and software engineering (e.g., Azure, Amazon Web Services); Healthcare | Data management and security | Top challenges for organisations undergoing digital innovation are costs, security, and expertise. | There is wide implementation of cloud, multi-cloud security tools and cost optimisation (FinOps) tools, and experimentation with AI/ML* and edge services. As a result, organisations report that governance, compliance and managing software licenses have become less of a challenge. | Y (Technological advances, Operational efficiencies, User focus, Functionality) | Y (User, Operational, Technological, Ethical) |
| Leong 2023 | Report | USA | Data science, IT and software engineering (e.g., Edge); Research and industry (e.g., research organisations) | Data management and security | Organisations’ IT infrastructures and data architectures are unfit for AI adoption, with data management and governance posing concerns due to skillset gaps, siloed working and issues around data control. | Cloud and Edge present a path to improving data architecture by accommodating AI/ML workloads, overcoming significant data challenges (e.g., source, type) and enabling data compliance and pre-processing for AI application. | Y (Technological advances, Operational efficiencies, Functionality) | Y (Operational, Technological) |
| Nexus FrontierTech 2023 | Report | China | Business, enterprise and finance; Data science, IT and software engineering (Nexus FrontierTech) | Data management and security; Data analytics and visualisation | Asset management faces data challenges such as response limitations, lack of systems for integrating internal data, and difficulties in mapping data to Environmental, Social and Governance (ESG) frameworks (central to asset management strategy). Adoption of AI/ML is lagging and mostly at the prototype stage. | A central Nexus platform comprising three AI models (ESSG Data Parser, News Monitoring, Investigative AI Models to standardise ESG integration within organisations, by managing ESG frameworks, data and news, allowing portfolio managers and analysts to visualise, benchmark and analyse company performance for impact investing. | Y (Technological advances, Operational efficiencies, User focus, Functionality) | Y (User, Operational, Technological) |
| Anonymous 2024a | Magazine | USA | Public administration, policy and legislature; Data science, IT and software engineering (WEKA); Business, enterprise and finance (KPMG) | Data management and security | AI practitioners identified data management as the primary technological inhibitor to AI/ML deployments, and data integrity as the top risk. | There is emphasis on educating others on the appropriate use of AI, focusing on valuable AI projects, and skills needs. | Y (Functionality) | Y (User, Operational, Ethical) |
| Nasir, Rizwan Ahmed and Bai 2023 | Preprint | Pakistan | Healthcare; Research and industry (DARPA) | Decision-making and strategy; Data management and security; Data analytics and visualisation | Questions around the explainability, accountability and transparency of AI hinder its adoption in clinical practice. | Development of Explainable Artificial Intelligence (XAI) techniques in healthcare are crucial. Techniques such as LIME and SHARP are commonly used and DARPA’s XAI program and toolkit is focused on development of an ethical framework that integrates governance, ethics and human oversight to regulate AI’s impact on individuals and society. | Y (Operational efficiencies, User focus, Functionality) | Y (User, Operational, Technological, Ethical) |
| Newman, Mintrom and O’Neill 2022 | Perspective | Australia | Public administration, policy and legislature | Data management and security; Data authentication | Public sector bureaucracy is being addressed with increasing computerisation and automation of services and administration. | AI has the potential to streamline bureaucratic processes deemed wasteful or obstructive to government action by performing human tasks faster and aiding in complex analysis and decision-making. If properly manged by humans, these technologies can also support neutrality and accountability in the public sector. | Y (Technological advances, User focus) | Y (Technological) |
| MIT Technology Review Insights 2022 | Report | USA | Data science, IT and software engineering (e.g., Databricks) | Data management and security; Data authentication | Embracing generative AI across sectors and industries requires strategic decisions on data infrastructure, model ownership, work force, and AI governance to fully harness the benefits and avoid losing competitive ground in the long term. | In preparation for generative AI, organisations are adopting flexible and scalable data infrastructures (e.g., data lakehouses) to democratice access to data and analytics and enhance data protection. Data lakehouses such as Dolly (Databricks) enable users to preform workloads and historical analysis with computational elasticity, while tailoring of generative models (e.g., BioBERT) allows domain-specific application. | Y (Technological advances, Operational efficiencies, User focus) | Y (Operational, Technological) |
| Anonymous 2024b | Magazine | USA | Data science, IT and software engineering (ePlus) | Data management and security | There is a significant gap in AI knowledge and readiness among organisations. | AI Ignite (ePlus) supports customer needs across various stages of readiness with solutions including workshops, readiness assessments, data strategy assessments, infrastructure builds, platform implementation and management, and ongoing support services | Y (Operational efficiencies, Functionality) | N |
| Nizami et al 2023 | Review | India | Healthcare | Data management and security; Data analytics and visualisation; Data authentication; Administration and communications; Decision-making and strategy; Research and insights | Big data analytics and ML are increasingly used for clinical data management to improve clinical trial outcomes and decision-making. | Integration of AI with electronic health records systems enables real-time data analysis, while predictive analytics helps identify at-risk patients, improve data quality by detecting errors and inconsistencies, and enhance patient care while reducing trial costs | Y (Operational efficiencies, Functionality) | Y (Operational, Technological, Ethical) |
| O’Keefe et al 2022 | Case study | USA | Business, enterprise and finance; Public administration, policy and legislature | Compliance and risk management | Financial institutions face increasingly stringent regulatory and internal demands, necessitating sophisticated risk management capabilities. | AI can aid managing and analysing large data volumes, but will require careful regulation (e.g., European Commission’s Artificial Intelligence Act 2021) evaluation, validating and human oversight to mitigate reputational, legal and regulatory risks (from improper use). | N | Y (User, Technological, Ethical) |
| Allan et al 2023 | Report | USA | Business, enterprise and finance; Research and industry (e.g., Sandia National Laboratories) | Data management and security | In a digital enterprise, strong and well-supported data infrastructures are crucial for seamless data flow – especially for workflows involving multiple institutions | Newer data infrastructures are designed to span multiple technological layers, facilitate AI/ML workflows and support data management and analysis – all while ensuring interoperability and FAIR data access across institutions | Y (Technological advances, Operational efficiencies, User focus, Functionality) | Y (User, Operational, Technological) |
| Ortega-Calvo et al 2023 | Journal article | Spain | Healthcare | Data management and security | Healthcare settings are in need of modern data platforms (MDPs) that can efficiently process and analyse large volumes of diverse, unstructured data. The challenges of building MDPs in this context is processing unstructured or heterogeneous data (e.g., electronic health records), ensuring user-friendliness, and ensuring management and governance of big data. | The Artifical intelligence Modern Data Platform (AIMDP) integrates MDP features with data science and ML capabilities (supervised and unsupervised learning) in a distributed Big Data environment | Y (Technological advances, Operational efficiencies, Functionality) | Y (Technological) |
| Owhonda et al 2022 | Journal article | Nigeria | Healthcare | Data management and security; Data analytics and visualisation; Data authentication | Data management platforms like SORMAS enable surveillance of disease outbreaks in resource-poor settings but require complex data entry (e.g., addresses), limiting the automation of data collection and analysis | A data analysis and validation engine (DAVE) was developed as a semi-autonomous data management solution for fast, automated data collection, processing and storage. The engine performs data quality control checks and uses ML to help classify data (e.g., addresses) | Y (Technological advances, Operational efficiencies, User focus) | Y (Operational) |
| Padarha 2023 | Preprint | USA | Other (generic) | Data management and security | Major tech (FAANG) companies have been involved in user privacy violations, raising concerns about unethical use of big data and biases in AI/ML systems. In particular, there is increased inequality around maths-based big data models. | Holding companies accountable and imposing tighter regulations is imperative for preventing the threat of big data models to democracies and human equality. | N | Y (Ethical) |
| Parthasarathy 2023 | Perspective | USA | Business, enterprise and finance; Healthcare | Data analytics and visualisation | With increased use of AI, collecting big data is becoming straightforward but interpreting it remains a challenge for enterprises. | Generative AI can facilitate data interpretation and quickly fill knowledge gaps by helping IT teams with observability, DevOps, and operations data analysis – reducing manual tasks and making data more accessibility. | Y (Technological advances, Operational efficiencies, User focus, Functionality) | N |
| Rajagopal et al 2022 | Conference proceeding | India | Food and Environment | Data management and security; Data analytics and visualisation; Decision-making and strategy | To stay profitable and make scientifically sound decisions, businesses must be able to effectively analyse and visualise large datasets. | AI-based decision support systems (DSS) can help inform decisions by analysing large datasets and presenting them graphically. | Y (Technological advances, Functionality) | N |
| Ramesh et al 2022 | Perspective | India | Healthcare | Data management and security | The ophthalmology healthcare sector needs new digital solutions for efficient sharing, management and analysis of big data. | Blockchain technology can assist AI algorithms in managing big data in ophthalmology, which is largely based on numerical data and imagery. It can also help monitor data and assess the integrity of analytical outputs. | Y (Technological advances, Operational efficiencies, User focus) | Y (User, Technological) |
| Rana et al 2022 | Report | India, Pakistan, Malaysia | Healthcare | Data management and security | Manual management of healthcare data (e.g., trial data, health records) is time-consuming and error-prone, limiting within-sector trust and collaborations. Efforts to improve this should focus on data interoperability, security and privacy. | Distributed ledger technology (e.g., Ethereum blockchain) can help manage digital assets in healthcare, ensure privacy and security of patient data and transparency of data exchange. | Y (Technological advances, Operational efficiencies, User focus) | Y (User, Operational) |
| Ravi et al 2022 | Perspective | USA | Research and industry (University of Illinois) | Data management and security; Data authentication; Research and insights | A common roadblock to using existing AI models for research is incompatibility of data, which hinders progress and discourages the adoption of AI methodologies. AI-ready data must be FAIR (findable, accessible, interoperable, reusable) | A proposed computational framework quantifies the FAIRness of AI models and trains them on AI-ready experimental datasets, enabling autonomous data management and AI-driven scientific discovery. | Y (Technological advances, Operational efficiencies, Functionality) | Y (Operational) |
| Raza et al 2024 | Preprint | Canada | Research and industry | Data management and security; Data authentication; Research and insights | Development of sophisticated large language models (LLMs) is challenging as it requires efficient data retrieval and analysis, interoperability of data formats and systems, and data accessibility | A proposed model development lifecycle for training LLMs incorporates FAIR data principles (Findable, Accessible, Interoperable, Reusable) at every stage and generates a FAIR-compliant dataset with diverse narratives | Y (Technological advances, Operational efficiencies, User focus) | Y (User, Operational, Technological, Ethical) |
| Reji et al 2023 | Journal article | Germany, Greece, Spain, Cyprus | Research and industry | Data management and security; Decision-making and strategy | Explainable AI (XAI) solutions can help ensure confidentiality and integrity of industrial data assets in manufacturing through access control, authentication, authorisation, regulation and auditing. | A proposed XAI platform for industrial asset handling and sharing ensures secure import, storage, transformation, management, sharing and tracking of industrial data – promoting automated production and manufacturing. | Y (Technological advances, Operational efficiencies, Functionality) | N |
| Riesener, Kuhn, Schuh 2022 | Report | Germany | Business, enterprise and finance; Data science, IT and software engineering | Data management and security; Data authentication | Data quality is a frequent barrier to companies’ digital transformation, requiring engineers to address issues such as low correctedness and low completeness to ensure reliable master data and harmonisation of datasets. | A four-step approach to enhancing master data management (MDM) uses AI to measure, assess and improve data quality. This improves the long-term efficiency of dataset management for enhanced business processes along the value chain. | Y (Technological advances, Operational efficiencies, User focus, Functionality) | Y (Ethical) |
| Rottkamp, Khan 2023 | Perspective | USA | Food and Environment | Data management and security; Data analytics and visualisation; Administration and communications; Decision-making and strategy | Combating climate change requires sustainable technology, whereby non-profits can effectively manage and analyse data long-term to target donors and identify opportunities and areas of operational improvement. | Sustainability can be achieved through automation of resource-intensive activities, predictive AI (e.g., impact of climate on business), advanced analytics for real-time performance analysis and cloud computing to enable remote work. | Y (Operational efficiencies, User focus) | N |
| Rousopoulou et al 2022 | Case study | Greece | Research and industry (European Commission) | Data management and security; Data analytics and visualisation; Decision-making and strategy | The challenges of the 4^th^ industrial revolution (Industry 4.0) lie in growth and complexity of technologies, as well as the customer demands for product quality and delivery time. Manufactures must balance product quality with efficiency and work with often very unstructured real-time data. | A proposed cognitive platform (supported by ML and deep learning) can autonomously deliver AI solutions and ensure quality results without needing constant monitoring and human resource allocation, thereby optimising factory management processes. | Y (Technological advances, Operational efficiencies, User focus, Functionality) | Y (User) |
| Roy 2022 | Preprint | USA | Research and industry (FAIR4HEP collaboration) | Data management and security; Research and insights | In recent years, digital object management practices have been adopted in data-intensive scientific disciplines to support findability, accessibility, interoperability, and reusability (FAIR) of research data | The FAIR4HEP collaboration promotes the FAIRification of data and AI models in higher energy physics (HEP), developing FAIRness metrics for experimental datasets and models, and providing an open-source notebooks for other HEP researchers to integrate FAIR principles into practice. | Y (technological advances, Operational efficiencies, User focus, Functionality) | N |
| Samuels 2023 | Perspective | USA | Data science, IT and software engineering (Snowflake) | Data management and security; Data analytics and visualisation | Workflows and AI models can vary significantly across business departments, making the jobs of data scientists, model developers and product owners (who must understand each workflow) more burdensome. | Automatically integrating governance and ethics into workflows and AI models will remove a lot of the burden and allow more focus on innovation. | Y (Operational efficiency, User focus) | Y (Technological, Ethical) |
| Hakem Al Harbi, Tidjon, Khomh 2023 | Preprint | Canada, Saudi Arabia | Data science, IT and software engineering (e.g., European Commission, IBM) | Data management and security; Data authentication; Decision-making and strategy | Ensuring responsible use of AI is becoming more crucial and should involve applying ethical principles throughout model development to ensure safe, fair and transparent business operations. | Numerous examples of ethical AI solutions include: the AI Ethics Orchestration and Automated Response (EOAR) framework for regular audits and testing of models, enabling comprehensive assessment of ethical compliance in AI systems; and Open Scale platform for defining, monitoring and tuning the ethical behaviour of IBM AI models | Y (Technological advances, Operational efficiencies, User focus, Functionality) | Y (User, Operational, Technological, Ethical) |
| Sen, Heim, Zhu 2022 | Review | USA | Data science, IT and software engineering | Data management | ML techniques are increasingly adopted for cybersecurity, but governments are concerned about the risks of adversarial AI. | AI-enabled intrusion detection and prevention systems (e.g., Darktrace Enterprise Immune System) can prevent cyberattacks, but holding their developers accountable means developing public policies and laws on AI cybersecurity. | Y (Technological advances, Operational efficiencies, User focus, Functionality) | Y (User, Operational, Technological, Ethical) |
| Serey et al 2023 | Review | Chile | Data science, IT and software engineering; Research and industry | Data management and security; Data analytics and visualisation; Research and insights | Managing large and asymmetrical collections of data presents ongoing challenges for companies, driving continuous development in big data management. | AI/ML-enabled automation, probabilistic reasoning and deep learning enable data analysis, pattern identification, anomaly detection, and predictive accuracy. | Y (Technological advances, Operational efficiencies) | Y (User, Operational) |
| Shaheen and Nemeth 2022 | Review | Hungary | Public administration, policy and legislature; Research and industry | Operations management and maintenance; Decision-making and strategy | In modern maintenance management practices, emphasis is placed on optimising costs and harnessing technology to deal with dynamic business environments. | Upgrading maintenance management systems to intelligent ones requires adopting big data, cloud computing and automation, developing cost-benefit models for investment predictions, managing vast data efficiently, addressing sustainability issues, and ensuring robust cybersecurity | N | Y (User, Operational) |
| Sheeder 2023 | Perspective | UK | Healthcare | Data management and security; Data analytics and visualisation; Administration and communications; Compliance and risk management | AI is improving efficiency in healthcare while also raising new challenges in data privacy, ethics and regulation – requiring a balance between advanced technology and human oversight | Aside from improving operational efficiency, predictive accuracy and cost-effectiveness, AI solutions must ensure integrity, confidentiality and stringent regulation of sensitive patient data. | Y (Technological advances, Operational efficiencies, Functionality) | Y (Operational, Technological, Ethical) |
| Shirvanian, Shams and Amir Masoud 2022 | Review | Iran, Taiwan | Data science, IT and software engineering | Data management and security | Traditional approaches to big data management are unsuitable for Internet of Things (IoT) environments, i.e., a network of interconnected and communicating devices. The goal is to create a seamless automated IoT environment where devices can exchange data efficiently and provide valuable data insights. | Solutions such as Edge computing reduce delays by processing data locally and exchanging only necessary data for analysis. Factors such as scalability, real-time data analysis, interoperability, and confidentiality are crucial in emerging IoT solutions (e.g., DeviceHive). | Y (Technological advances, Operational efficiencies, User focus, Functionality) | Y (User) |
| Simonofski et al 2022 | Journal article | Belgium, Netherlands | Public administration and legislature | Data management and security; Data analytics and visualisation; Compliance and risk management | As the use of AI analytics grows, public administrations must balance the technological benefits with compliance to legal requirements like GDPR. | Balancing advanced analytics with legal requirements necessitates proportionate data collection and use, timely access to only the necessary data, and being transparent with citizens about all data operations. | N | Y (User, Technological, Ethical) |
| Souza 2022 | Perspective | UK | Business, enterprise and finance | Compliance and risk management | AI brings significant benefit to finance, but its regulation is fragmented, posing challenges for institutions in addressing the AI risks. | Regulatory efforts are underway to enhance existing AI model risk management frameworks, particularly in terms of governance, ethics and lifecycle updates, with better guidance for mitigating risks. | Y (Technological advances) | Y (Technological, Ethical) |
| Srivastava 2023 | Journal article | Bihar | Other (generic) | Data management and security; Data authentication; Decision-making and strategy | AI is revolutionising data management, integration and analytics through automation, predictive capabilities and adaptive learning, providing more informed strategies for decision-makers. | AI/NLP (natural language processing) offers particular benefits to data processes, such as automated data collection, error detection, continuous monitoring and auditing for enhanced insights and decision-making. | Y (Technological advances, Operational efficiencies, User focus, Functionality) | N |
| Stoitsis and Manouselis 2023 | Journal article | Greece | Food and Environment; Data science, IT and software engineering | Data analytics and visualisation | AI technology can be deployed to predict food risks and prevent food safety incidents, reducing unsustainable and costly food recalls. | Numerous open-source initiatives and frameworks (e.g., Apache Spark) are using AI to enhance food safety systems, creating software for monitoring supplier compliance, scanning food incidents and addressing broader food safety challenges. | Y (Technological advances, Operational efficiencies, User focus, Functionality) | N |
| Stoykova and Shakev 2023 | Review | Bulgaria | Data science, IT and software engineering | Data management and security | The capabilities of AI can be utilised to enhance management information systems and increase business value and competitiveness. | Process automation enables humans to focus on higher-value activities, while ML and deep learning (e.g., GPT-4) can speed up data analysis and enhance predictive insights. AI assistants (e.g., Conversational AI) can enhance customer experiences and support corporate onboarding and learning. | Y (Technological advances, Operational efficiencies, User focus, Functionality) | Y (User, Operational, Technological, Ethical) |
| Sung-Chu 2023 | Webpage | USA | Business, enterprise and finance | Data management and security; Administration and communications | Empathy is becoming more important in organisational health, financial outcomes and employee wellbeing. | Empathetic AI is being increasingly integrated into and enhancing efficiency in organisational processes, allowing leadership and HR to focus more on supporting employees (e.g., with meaningful feedback) through automation. | Y (Technological advances, Operational efficiencies, User focus, Functionality) | Y (User, Operational, Technological, Technological) |
| Tandale et al 2022 | Perspective | India | Public administration, policy and legislature; Data science, IT and software engineering; Business, enterprise and finance | Data management and security; Data analytics and visualisation | Despite the growing use of AI, there is limited research on how entrepreneurial orientation (EO) influences its adoption and impact – including direct and indirect effects on operating efficiency. | Many organisations are still in the data gathering stage of building their AI strategy. It is believed that AI can aid organisational staff in analytics and decision-making with intelligent tools (e.g., cognitive computing) and executive mechanisms, transforming consumer experiences and increasing return on investment. | Y (Technological advances, Operational efficiencies, User focus, Functionality) | Y (Operational, Technological) |
| Tang et al 2023 | Preprint | China, USA | Data science, IT and software engineering | Data management and security; Decision-making and strategy | Generative AI is being increasingly used, but its outputs should be verified by analysing supporting data to ensure accuracy, quality and consistency for reliable decision-making. | VerifAI is as a proposed modular framework for verifying the reliability of AI by analysing supporting data from multi-modal data lakes. Experiments show it achieves high accuracy and is extendable to different data types and sources. | Y (Technological advances, Operational efficiencies, User focus, Functionality) | Y (Operational, Technological, Ethical) |
| Tito 2023 | Thesis | Finland | Research and industry; Business, enterprise and finance | Data management and security; Data authentication | The rise of digital transformation, advanced analytics and data driven-decision-making necessitates a shift towards comprehensive more data management practices, with focus on data quality and its value add to business efficiency. | Data warehousing and cloud computing offer solutions for big data management and decision support, enabling comprehensive governance frameworks that enhance data quality, efficiency and security in preparation for business growth. | Y (Technological advances, Operational efficiencies, User focus, Functionality) | Y (User, Operational, Technological, Ethical) |
| Valle-Cruz and García-Contreras 2023 | Journal article | Mexico | Public administration, policy and legislature | Data management and security | The challenges with smart data management and AI in the public sector include addressing data privacy concerns, navigating ethical risks and ensuring the readiness of public sector employees to effectively utilise technologies. | Successful AI implementation in the public sector requires embracing technological change, understanding its benefits (e.g., providing efficient and scalable computing environments during Covid-19) and replacing outdated systems and leveraging data to meet citizen needs effectively | Y (Technological advances, Operational efficiencies, User focus, Functionality) | Y (Technological, Ethical) |
| van Noordt, Medaglia, Tangi 2023 | Journal article | Estonia, Denmark, Italy | Public administration, policy and legislature | Data management and security; Decision-making and strategy | EU national policy initiatives aim to foster AI development and adoption in public administrations, addressing common barriers (e.g., data quality, ethics, AI expertise interorganisational collaboration) and enhancing social benefit and economic growth. | AI can be harnessed to address the needs of citizens, identify emerging societal problems and ensure decision-making is targeted to the wellbeing of citizens. However, this would require organisations and administrations to replace old systems with technologies enabling automation and efficient workflows. | Y (Operational efficiencies, User focus, Functionality) | Y (Technological, Ethical) |
| van Noordt and Misuraca 2022a | Journal article | Estonia, Italy | Public administration, policy and legislature | Data management and security; Decision-making and strategy | AI is widely used in the EU but little evidence of its effect on core governance functions, policy and public sector impact exists. | The integration of big data and AI into public administration can enhance and streamline identifying and solving citizens’ problems, automate routine tasks, improve resource allocation and increase public service delivery efficiency. | Y (Technological advances, Operational efficiencies, User focus, Functionality) | Y (User, Operational, Technological, Ethical) |
| van Noordt and Misuraca 2022b | Journal article | Estonia, Austria | Public administration, policy and legislature | Administration and communications | Ensuring effective adoption of AI in EU’s public administrations requires understanding the antecedents (e.g., historical barriers) of public sector innovation to better understand the conditions under which AI innovations can develop. | The potential barriers to adopting AI include accountability (e.g., ‘black-box’ decision-making), privacy risks, discrimination and bias, inadequate resource, infrastructure and expertise. | Y (Technological advances, Operational efficiencies) | Y (User, Operational) |
| van Noordt and Misuraca 2023 | Journal article | Estonia, Italy | Public administration, policy and legislature | Administration and communications | There is limited empirical evidence on what constitutes AI capability, how it can be acquired and what factors confer/affect the integration and implementation of AI in public administrations. | Developing public AI capability is closely tied to 1) digitalisation efforts built on past eGovernment initiatives, but require new data and adherence to ethical standards, 2) internal or externally sourced infrastructure and continuous training, and 3) stable financial resources for maintaining and scaling AI systems. | Y (Operational efficiencies, User focus, Functionality) | Y (User, Operational, Technological, Ethical) |
| Priya et al 2023 | Journal article | India | Business, enterprise and finance | Data management and security; Data analytics and visualisation; Operations management | The evidence gap in AI’s role in enhancing data integrity and security across sectors needs to be addressed, with a focus on the potential challenges to AI adoption. | While AI distinguishes itself from traditional analytics tools by automating data processing and transforming the impact of existing applications, there are crucial considerations related to ensuring that the usage of data aligns with its intended purposes. | Y (Operational efficiencies, User focus, Functionality) | Y (User, Operational, Technological, Ethical) |
| Vrabie 2022 | Conference proceeding | Romania | Public administration, policy and legislature; Data science, IT and software engineering (e.g., Google, Apple) | Data management and security; Administration and communications | There is potential for AI to enhance operations and service delivery in smart cities and governments, transforming them from reactive to proactive. Policymakers require insight into AI solutions and how to integrate these into strategy amid budget constraints and declining trust in public services. | AI can streamline public administration in numerous ways, such as through automation (e.g., Robotic Process Automation), regulation, enhanced e-procurement and insightful reports (e.g., on budgets, citizen requests), with the end goal of improving efficiencies, reducing redundancies and increasing public confidence in services. | Y (Technological advances, Operational efficiencies, User focus, Functionality) | Y (Technological, Ethical) |
| Weber et al 2023 | Journal article | Germany, Australia | Business, enterprise and finance | Data management and security | Organisations should develop specific capabilities for implementing AI, particularly in understanding how these facilitate coping with AI’s unique characteristics, such as inscrutability and data dependency. | Organisational capabilities could be categorised into 4 key areas: 1) AI Project Planning, 2) Co-Development for managing project complexity and stakeholders, 3) Data management, and 4) AI Model Lifecycle Management for addressing data challenges and evolving AI systems. | Y (Operational efficiencies, User focus) | Y (User, Operational, Technological, Ethical) |
| Weinert et al 2022 | Journal article | Germany | Healthcare | Data management and security | Despite the rapid development of AI, strategies and practical experiences for adopting AI in healthcare are lacking. The complexity of AI can therefore reduce trust among decision-makers and users in healthcare. | In order to deal with the challenges, including lack of resources, compatibility, data quality and training costs, healthcare organisations must develop competencies, workflows and address technical requirements (e.g., hardware, software, interoperability). | Y (Operational efficiencies, User focus) | Y (User, Operational, Technological, Ethical) |
| Whang et al 2023 | Journal article | South Korea | Data science, IT and software engineering (e.g., Google) | Data management and security; Data authentication | Research institutions frequently allocate 90% of their ML efforts to algorithms and only 10% to data pre-processing, although it should be the reverse. Data-centric AI efforts focus on improving data pre-processing and therefore model accuracy. | Tools and techniques for data discovery, augmentation, validation and cleaning in ML include Goods (Google’s data lake) Jupyter Notebook (interactive data management), AutoAugment (automates data transformation) and Mixup (data augmentation). | Y (Technological advances, Operational efficiencies) | Y (Operational, Technological) |
| Williamson et al 2023 | Perspective | UK | Research and industry | Data management and security; Data authentication; Research and insights | A major barrier in developing reliable AI is creating effective data management strategies that integrate large, multi-dimensional datasets from different scientific disciplines (e.g., as in agricultural technology). | Possible solutions to data challenges include: adopting FAIR principles (making data findable, accessible, interoperable and reusable); providing guidance on AI data requirements; documenting material provenance; sharing semantic standards; data quality benchmarking; open and adaptable models; open access to data; investment in data services | Y (Technological advances, Operational efficiencies, User focus) | Y (Operational, Technological, Ethical) |
| Wróbel 2022 | Perspective | Poland | Public administration, enterprise and legislature | Data management and security; Administration and communications | For the use of AI in public administrations in the EU to become widespread, it needs to be assessed whether the EU laws underlying traditional administration systems support AI adoption or whether they must be changed. | A clear AI definition is essential for an effective legal framework to ensure individuals can fully exercise their right to good administration. Current proposals however lack this, posing various risks, and EU institutions must therefore clarify citizens’ administrative rights and authorities’ obligations when regulating AI. | N | Y (User, Technological, Ethical) |
| Yamani, Alsunaidi, Boudellioua 2022 | Journal article | Saudi Arabia | Data science, IT and software engineering (e.g., Microsoft); Research and industry | Data management and security | ML techniques depend on the volume and diversity of training data, however the problem that needs to be addressed is how big data can be processed and where to store it. | There are numerous software tools that provide storage and distributed processing for big data, such as Hadoop, Microsoft Azure and Spark. The most popular, Apache Spark, is an open-source platform that supports several programming languages and includes many capabilities. | Y (Technological, User focus) | N |
| Yang et al 2024 | Journal article | China, USA | Healthcare; Data science, IT and software engineering (e.g., Apache) | Data management and security; Data authentication | Biomedical researchers developing precision medicine face significant challenges in curating large volumes of diverse digital data, hampered by high technology costs and limited access to big data tools. | Precision medicine can benefit from numerous big data platforms (e.g., Apache Hadoop, Spark), databases (e.g., MongoDB), and data integration tools (e.g., Health IT) for managing and analysing large-scale genomic and clinical datasets, integrating bioinformatics with data science. | Y (Technological advances, Operational efficiencies, User focus, Functionality) | Y (User, Operational, Technological, Ethical) |
| Yu et al 2023 | Journal article | Australia, USA | Healthcare; Data science, IT and software engineering (e.g., Google) | Data management and security; Administration and communications | Generative AI and large language models (LLMs) hold significant promise for healthcare, but also present critical limitations and ethical concerns – requiring a strategic roadmap for their effective healthcare integration, applications, techniques, and regulatory considerations. | The 3 critical factors for selecting LLMs (e.g., Google’s PaLM) for healthcare are models, data and tasks. These favour instruction fine-tuned LLMs for their contextual understanding, alignment with specific applications, high-quality and diverse training data, and enhanced performance, with potential benefits including reducing clinical documentation burden, better interpretability and hence health outcomes. | Y (Technological advances, Operational efficiencies, User focus, Functionality) | Y (User, Operational, Technological, Ethical) |
| Yu, Zhang and Wang 2022 | Journal article | China | Healthcare | Data management and security; Administration and communications | Service-oriented architecture offers a flexible design for deploying discrete AI models, but lacks specialised development language support, posing challenges for hospital human resource management systems; these handle diverse data types and must ensure both internal and external access while providing effective data visualisation for scientific insights. | To address healthcare needs using AI, it is crucial to select appropriate models, use specialised, high-quality pre-training data and apply models to tasks like natural language understanding and generation, while employing learning from human feedback to improve clinical workflows and data management. | Y (Technological advances, Operational efficiencies, User focus, Functionality) | Y (User, Operational, Technological, Ethical) |
| Zárate et al 2022 | Journal article | Spain | Research and industry | Data management and security; Data analytics and visualisation | In ML operations, an experimentation catalogue that tracks different experiments and data is essential for managing and reproducing AI processes. This can be achieved by integrating data and model catalogues, supported by automated infrastructure and versioning tools for seamless dataset and AI process management. | The creation of data and model catalogues with Knowledge to Environment (K2E) aims to address challenges with existing tools (e.g., DVC), which often have limitations in dataset versioning, ease of use, storage technology support, and reproducibility, making them reliant on the practices of data scientists. | Y (Technological advances, Operational efficiencies, User focus, Functionality) | Y (Technological) |
| Zhang et al 2023 | Preprint | China | Business, enterprise and finance | Data management and security; Data analytics and visualisation | Large language models (LLMs) like ChatGPT and GPT-4 face significant challenges in managing, processing and displaying large-scale data due to limitations in direct data reading, numerical computations, external tool integration, and diverse data-related tasks requiring diverse output formats. | Data-Copilot is a universal framework that uses LLMs to autonomously manage, process and visualise diverse datasets according to user preferences. This significantly reduces the need for manual effort and expert knowledge by acting as an AI assistant for complex data tasks. | Y (Technological advances, Operational efficiencies, User focus, Functionality) | Y (Operational, Technological) |
| Anonymous 2022c | Magazine | USA | Data science, IT and software engineering (e.g., Snowflake) | Data management and security; Data analytics and visualisation | IT experts are struggling with data management (capturing, organising, storing, searching, transferring) and improving data quality for better analytics and business insights. | Investing in automated and time-saving tools (e.g., metadata management platforms) can significantly improve the accuracy and efficiency of data operations, while modern data architectures (e.g., data lakehouse, fabric/mesh) can help integrate data cross multi-cloud environments and help handle complex, hybrid data | Y (Technological advances, Operational efficiencies, User focus, Functionality) | N |
| Chen et al 2024 | Preprint | USA | Research and industry (Carnegie Mellon University) | Data management and security | Development and implementation of AI/ML entails repetitive tasks (ML practitioners must train and evaluate models and test their performance) and productivity is hampered by the manual labour involved in the data management, coding and record-keeping needed for model reproducibility. | Accelerated Cloud for AI (ACAI) offers ML practitioners a data lake for storing versioned datasets with metadata and an execution engine for running ML jobs on the cloud, with automatic resource provisioning, logging and provenance tracking. This results in a 1.7x speed-up, 39% cost reduction and a 20% reduction in experiment time (for typical ML use cases). | Y (Technological advances, Operational efficiencies, User focus, Functionality) | Y (User, Operational) |
| Medina et al 2022 | Perspective | Qatar, Norway, Denmark | Research and industry (e.g., Hamad Bin Khalifa University) | Data management and security; Data authentication | Good research data management is crucial, but increasing use of bid data demands efforts to ensure data is FAIR (findable, accessible, interoperable, and reusable) and addressing the ‘5V challenge in big data’ (volume, variety, velocity, veracity, value) to ensure data quality, accuracy and good ML model performance. | Data pre-processing should be an essential part of the research lifecycle, and include data cleaning, normalisation, and feature extraction. Tools such as AiiDA (an AI-powered digital assistant) can automate and centralise data flow, collection and dissemination, and is tailored for researchers specifically. | Y (Technological advances, Operational efficiencies, User focus, Functionality) | N |
| Konovalova 2023 | Perspective | Ukraine | Public administration, policy and legislature | Data management and security; Decision-making and strategy | Promoting international cooperation in digital technologies, including AI, is diplomatically crucial for maintaining democracy and geopolitical relevance. Key challenges include ensuring AI is deemed lawful, ethical and socially reliable. | AI can be useful for diplomatic messages – distinguishing genuine messages from background noise. However, there is concern for the diplomacy profession, that it may be replaced by machines, and existing diplomatic services will have to be restructured to catch up with the speed of digital and AI developments. | Y (Functionality) | Y (User, Ethical) |
| Chen, Wu and Wang 2023 | Review | China | Research and industry; Data science, IT and software engineering (e.g., IBM) | Data management and security; Decision-making and strategy | Human bias infiltrates AI-driven decision-making due to human involvement in data collection and ML training. Biases in ML can therefore arise from unawareness, incomplete data or annotation errors and can potentially prejudice AI decisions against certain social groups. | AI fairness can be enabled by developing fairness metrics (e.g., disparate impact, equal opportunity, statistical parity), mitigating algorithmic bias (e.g., through training) and fair representation learning frameworks. Existing tools (e.g., IBM AI Fairness 360) can help detect and mitigate bias in AI models. | Y (Technological advances, Operational efficiencies, User focus, Functionality) | Y (User, Operational, Ethical) |
| Chowdhury 2023b | Perspective | India | Other (education) | Decision-making and strategy; Administration and communications | Post-Covid, the emphasis on digital transformation in academia aims to streamline complex internal educational administration processes, such as planning, organising, managing resources and staffing, evaluating, and coordinating activities. | AI-driven scheduling software and intelligent assistants can reduce administrative burden, however unethical behaviour around AI use in educational settings is on the rise; moreover, there is little knowledge and resource to support AI in management, and not all staff are willing to adopt new digital skills. | Y (Technological advances, Operational efficiencies, Functionality) | Y (User, Operational, Technological, Ethical) |
| Manias et al 2023 | Web page | European Union | Public administration, policy and legislature (European Research Agency and its international consortia) | Decision-making and strategy | AI can be publicly harnessed to solve the numerous challenges of modern democracies like the EU, but guidance, support and regulation are needed. | The EU’s AI4GOV project aims to design a reference framework for ethical and democratic AI use, develop AI fairness monitoring (and bias mitigation), create trusted AI techniques for AI explainability (for policymakers, citizens etc), and train stakeholders and citizens in democratic AI elements to support democratic processes and boost regulatory compliance (e.g., with GDPR). | Y (Technological advances, Operational efficiencies, User focus, Functionality) | N |
| Benito 2024 | Magazine | Turkey | Business, enterprise and finance; Data science, IT and software engineering | Data management and security; Data analytics and visualisation; Administration and communications | To establish a strong digital core for future AI readiness, organisations must renovate outdated foundations, harmonise business processes, unify and standardise master data and infrastructure, and enhance IT agility, competitiveness and resilience. | Cloud-based solutions (e.g., Cloudera, Azure cognitive services) can support humans in routine tasks, shorten the time taken to prepare corporate reports, enable quicker strategic decisions, and support the democratisation of data. | Y (Technological advances, Operational efficiencies, User focus, Functionality) | Y (User, Operational, Technological) |
| Boban and Klaric 2023 | Magazine | Croatia | Public administration, policy and legislature; Healthcare | Data management and security; Decision-making and strategy | A key aim for European AI initiatives is to foster trust and responsibility, ensuring GDPR compliance and provisions for automated decision-making – particularly in healthcare. | The EU AI Guidelines provide an assessment list to ensure AI systems continuously meet criteria for transparency, privacy and data governance, technical robustness and safety, human agency and oversight, accountability, societal and environmental wellbeing, and diversity and fairness. | N | Y (User, Operational, Technological, Ethical) |
| Holmes and Douglass 2022 | Journal article | USA | Healthcare | Data management and security | The digital skills gap poses a significant threat to the workforce, with external regulators struggling to keep pace with AI advancements. This places the burden of AI implementation on organisations, who must ensure they have the expertise and resource to ensure control of their AI systems. | Two key areas for automation in accounting are Robotic Process Automation, which automates repetitive tasks, and ML, which utilises deep learning for predictive performance and audit support, while also providing evidential reasoning for improved risk assessment and audit quality assurance. | Y (Technological advances, Operational efficiencies) | Y (User, Operational, Technological, Ethical) |
| Linthicum 2023 | Magazine | USA | Data science, IT and software engineering | Data management and security | Despite the clear benefits of generative AI in the cloud (e.g., scalability, cost-efficiency), its widespread adoption is hampered by a significant skills gap, as implementing these models requires advanced expertise in ML, cloud computing and data engineering. | Organisations must invest time and resource to ensure data availability and quality, carefully evaluate cloud resources for model training, and address the skills gap to successfully implement generative AI models in the cloud. | N | Y (User, Operational, Technological, Ethical) |
| Agrawal, Sahu and Kumar 2022 | Review | India | Public administration, policy and legislature | Data management and security; Data analytics and visualisation; Decision-making and strategy | Unstructured data poses challenges for traditional databases used in legal informatics systems, necessitating tools to automate repetitive tasks and allow more focus on valuable activities. | Digital transformation of legal environments and integration of advanced offers benefits such as real-time monitoring, data transparency, and cost-effectiveness, with AI-based systems and structured data platforms improving decision-making and predictive analytics. | Y (Technological advances, Operational efficiencies, User focus, Functionality) | Y (User, Operational, Technological, Ethical) |
| Choi et al 2023 | Journal article | Korea | Healthcare | Data management and security; Administration and communications | Healthcare data is rapidly growing and needs to be managed, however big data management requires addressing cost and monitoring issues, establishing stable protocols, and navigating various data governance regulations. Moreover, AI applications must be integrated without disrupting clinical systems. | A ‘digital twin’ of real-world data, a viable healthcare solution, encompasses people, hospitals, systems and diseases, enabling safe and effective virtual reality simulations (e.g., training, education, remote medical consultations) and medical AI services. | Y (Technological advances, Operational efficiencies, User focus, Functionality) | Y (User, Operational, Technological, Ethical) |
| McKendrick 2023 | Magazine | USA | Data science, IT and software engineering | Data management and security | In recent years, there has been a significant trend of migrating data to the Cloud, but this has introduced some complexities. Issues such as data duplication and challenges around data governance, security, storage and quality management have emerged. | Data mesh and data fabric are emerging as next-generation architecture solutions to data management issues. Built on Cloud, these platforms are scalable, enhance data discovery, streamline data flows and enable real-time insights for business performance. | Y (Technological advances, Operational efficiencies, User focus, Functionality) | Y (User, Operational, Technological, Ethical) |
| Dunleavy and Margetts 2023 | Review | UK | Public administration, policy and legislature | Administration and communications; Decision-making and strategy | Governments are shifting from ‘digital governance’ to ‘algorithmic governance’, but still trail the private sector’s advancements – with AI still being perceived as a niche area in public administrations. | The ’Digital-Era Governance’ model proposes a framework for analysing how digitalisation impacts administrations, themed around 1) capability to store and analyse big data, 2) capability to extend workers’ capabilities, 3) maximising organisational productivity and 4) administrative holism. | Y (Technological advances, Operational efficiencies, User focus, Functionality) | Y (User, Operational, Technological, Ethical) |
| Anonymous 2022b | Magazine | USA | Data science, IT and software engineering (e.g., Snowflake) | Data management and security | As ML adoption becomes standard in the data industry, the limitations of current data structures are evident in their inefficiency. Scaling data models across organisations introduces complexities, reduces productivity and hinders efficient data processing. | ML experts are devising strategies to enhance data value and support enterprise growth. Snowflake’s Snowpark is an example of a solution that integrates diverse code languages and data model tools, streamlining data pipelines and governance for ML-enabled automation. | Y (Technological advances, Operational efficiencies, User focus, Functionality) | Y (User, Operational, Technological, Ethical) |
| Bentum 2023 | PhD thesis | USA | Research and industry | Data management and security; Data authentication | Replicating the AI success of private sectors in research has been a challenge due to difficulties in achieving AI-ready data – i.e., data that meets FAIR criteria (findability, accessibility, interoperability, reusability) for effective AI model training and performance. Moreover, there is lack of expertise, with AI consultancy posing a costly, unscalable option. | Organisations have started engaging with data science teams to post-process their datasets, who manually or with the help of software clean, assemble and validate the data. Where AI is experimentally used on imperfect data, this reduces some processing effort and guides future data collection practices. | Y (Technological advances, Operational efficiencies, User focus, Functionality) | Y (User, Operational, Technological, Ethical) |
| Bobrovskyi et al 2023 | Review | Ukraine | Research and industry; Public administration, policy, and legislature | Administration and communications | The digitalisation of public administrations and research environments will centre around achieving efficient and secure digital public services, unifying disparate data across national and regional administrative bodies. There is particular focus on transforming HR systems. | HR transformation can be achieved with neural networks, AI, natural language processing (NLP) and gamification, enabling Cloud-based systems to be fully automated (e.g., through ML) and offering tools such as recruiter robots, chatbots (for administrative tasks), cognitive selection and digital referral recruiting platforms. | Y (Technological advances, Operational efficiencies, User focus, Functionality) | N |
| Gaur 2023 | Magazine | India | Data science, IT and software engineering (Hitachi) | Data management and security | Successful AI implementation requires precise and dependable, however only 3% of companies have data that adheres to fundamental quality standards (according to a Harvard Business Review). | Pentaho+ (available through Hitachi/AWS/Azure marketplaces) supports robust, simplified data management and operations using precise data that can be integrated with existing infrastructures and used for big data analytics and business intelligence. | Y (Technological advances, Operational efficiencies, User focus, Functionality) | N |
| Huerta et al 2023 | Perspective | USA, Greece, Germany, Sweden, Finland | Research and industry (e.g., National Science Foundation) | Data management and security; Data authentication | Ensuring research data complies with FAIR principles (findability, accessibility, interoperability, reusability) requires investing into digital assets, like research software, workflows and AI models. | Emerging research-driven initiatives in the US and Europe (e.g., HPC-FAIR) aim to redefine and apply FAIR principles to maximise the impact of data investments and promote innovation. Projects include testing of FAIR metrics, simulation-based research, and enabling ethical inquiries into datasets. | Y (Technological advances, Operational efficiencies, User focus, Functionality) | Y (User, Operational, Technological, Ethical) |
| Hasnain 2024 | Blog | USA | Business, enterprise and finance; Data science, IT and software engineering | Data management and security | In the era of big data, data sets are so vast that humans are unable to manually analyse them, hindering the extraction of valuable insights for business intelligence. | AI is transforming software development and enhancing user experiences by automating tasks (e.g., Astera’s data management platform) and streamlining data pipelines; however, this requires organisational buy-in, data literacy and alignment of AI efforts with business goals. | Y (Technological advances, Operational efficiencies, Functionality) | Y (User, Operational, Technological) |
| Mahmood et al 2023 | Journal article | United Arab Emirates | Business, enterprise and finance | Administration and communications | With the increasing complexity of businesses, traditional, traditional techniques and tools for project management are proving inadequate. Project managers should therefore leverage AI to track project workflows and use real-time information on schedule, costs and risks for decision-making. | AI can enhance project management through automation and intelligent decision-making (e.g., predicting costs, risks), increasing productivity and allowing for more complex problems to be solved. Example tools are knowledge based expert systems, fuzzy logic, and artificial neural networks. | Y (Technological advances, Operational efficiencies, User focus, Functionality) | Y (User, Operational) |
| Cho, Choi and Choi 2023 | Review | New Zealand | Data science, IT and software engineering (e.g., IBM); Public administration, policy and legislature | Data analytics and visualisation; Administration and communications | In growingly complex business environments, personnel management and HR decisions can no longer solely rely on managers’ intuition and experience. HR analytics has emerged as a modern trend in public and private organisations. | HR analytics leverages advanced technologies like 5G and AI (e.g., IBM’s virtual assistant platform, Your Learning) to streamline training, talent planning and recruitment processes, enhancing performance and employee satisfaction across public and private sectors. | Y (Technological advances, Operational efficiencies, User focus, Functionality) | Y (User, Operational, Technological, Ethical) |
| Cavanagh, Pariona-Cabrera and Halvorsen 2023 | Perspective | Australia | Healthcare | Data management and security; Data analytics and visualisation; Administration and communications | HR managers in healthcare experience burnout from applying outdated HR processes to increasingly complex service issues. Efforts to enhance staff performance and organisational effectiveness underscore the need for HR analytics to monitor staff wellbeing and improve performance. | AI in HR management can enhance recruitment, operations, analytics and reporting, while offering self-service assistance, learning and development. It can also support clinicians with data analysis to improve patient care quality and clinician productivity, in all giving staff more job autonomy and satisfaction (e.g., by allowing them to focus on vital tasks and patient care). | Y (Technological advances, Operational efficiencies, User focus, Functionality) | N |
| Chowdhury 2023a | Book | USA | Research and industry | Data management and security; Data analytics and visualisation; Decision-making and strategy | To remain competitive and sustainable in a tech-driven environment, and ensure customer retention, organisations should adopt AI/ML to harness their data assets for business outcomes. | Data mining algorithms and artificial neural networks can inform a business intelligence (BI) model that enables organisations to seize market opportunities and counter potential threats. The three-tier model is built on: 1) a data warehouse (Snowflake), 2) a data extraction model (ETL) and 3) BI report generation. | Y (Technological advances, Operational efficiencies, User focus, Functionality) | N |
| Foffano, Scantamburlo and Cortés 2023 | Journal article | USA, Italy, Spain | Public administration, policy and legislature (EU member states) | Decision-making and strategy | EU member states are working to harness AI for societal benefit through national AI strategies, aiming to balance innovation with ethical sustainability. To achieve this, the European Commission has introduced initiatives such as policy documents, ethics guidelines, and a proposal for regulation. | Framing AI for social good involves focus on 1) applications, 2) ethical principles, and 3) policies. EC’s initiatives aim to boost technological and industrial capacity; prepare for socioeconomic changes; and ensure an ethical and legal framework. | Y (Technological advances, Operational efficiencies, Functionality) | Y (Ethical) |
| Bracci 2022 | Review | Italy | Public administration, policy and legislature; Other | Decision-making and strategy | There are concerns of “increasing blurring of accountabilities” in digitalised governments. Although accountability in AI remains an ethical issue, only recently have scholars begun to focus on its implications for public services. | Strengthening accountability in AI (‘explainable AI’) involves increasing oversight and human involvement through the model lifecycle, engaging internal stakeholders and AI users to avoid automation bias. | Y (Technological advances, Operational efficiencies, User focus, Functionality) | Y (User, Operational, Technological, Ethical) |
| McKendrick 2022 | Perspective | UK | Data science, IT and software engineering | Data management and security | Challenges in cloud data governance that need to be addressed include integrating data across hybrid environments; increasing data security; expanding data management; and achieving greater observability in the data pipeline. Moreover, AI/ML require robust data processing to ensure accuracy. | Recommendations for enterprises using cloud data governance for AI include: automating the governance process, not outsourcing data governance, aligning it with business, understanding who creates and consumes data, and identifying key internal partners. | Y (Technological advances, Operational efficiencies) | Y (User, Operational, Technological, Ethical) |
| Informatica 2024 | Report | USA | Data science, IT and software engineering | Data management and security; Data authentication | Data leaders are capitalising on generative AI and considering upskilling their employees in AI/ML. Most foresee increased interest and investment in AI and data management solutions. | The key metrics of an effective data strategy are: improved data readiness for AI, improved data literacy, improved data quality and increased data management and analytics efficiency. | Y (Technological advances, Operational efficiency, Functionality) | Y (Technological, Ethical) |
| Harrer 2023 | Perspective | Australia | Healthcare | Data management and security | Without human oversight, generative AI applications risk being perceived as mere novelty tricks and spreading misinformation or harmful content. There is also concerns about the unsafe deployment and exaggerated expectations of large language models (LLMs) in healthcare. | The WHO’s 2021 framework outlines ethical principles for integrating LLMs into healthcare AI systems, emphasising accountability, fairness, data privacy, transparency and alignment with values. With this, LLMs have the potential to streamline documentation, improve clinical trial efficiency and enhance interpretation of electronic health records. | Y (Technological advances, Operational efficiency, User focus, Functionality) | Y (User, Operational) |
| eClinical Solutions 2024 | Report | USA | Healthcare | Research and insights; Data analytics and visualisation | The increasing complexity of clinical, associated with more voluminous and diverse data, has led to increased interest in AI/ML and how these tools could transform not only trial data management, but design, risk assessment and analysis. | eClinical Solutions provides a leading data/analytics platform with biometrics services that enables researchers to manage trials efficiently, gain timely insights and scale resulting innovations using a scalable AI-integrated clinical data infrastructure. | Y (Technological advances, Operational efficiencies) | Y (User, Operational) |
| Hradecky, Kennell and Davidson 2022 | Journal article | UK | Business, enterprise and finance | Administration and communications | Unlike other digital technologies, the unique characteristics of AI present a significant knowledge barrier to organisations’ leaders, staff and technical teams. | Tornatzky and Fleishcher’s (1990) Technology-Organization-Environment (TOE) framework is suggested as a useful tool in organisational decision-making for considering the technological, organisational and environmental factors when adopting new technologies. | Y (Technological advances, Operational efficiencies, User focus) | Y (User, Operational, Technological) |
| Katirai et al 2023 | Journal article | Japan | Healthcare | Administration and communications | There is a patient and public involvement (PPI) gap when it comes to AI in healthcare. While patients and members of the public are the end users of healthcare, little is known about their views on the use of AI to make healthcare decisions. | Japan and UK’s AIDE Project exemplifies a collaborative PPI exercise, whereby an exploratory PPI Panel (PPIP) of patients, caregivers and the public was used to understand expectations and concerns around the use of AI in healthcare (e.g., changes in roles and relationships, quality of care). | Y (Technological advances, Operational efficiencies, User focus, Functionality) | Y (User, Operational, Technological, Ethical) |
| Gamble 2023 | Perspective | USA | Business, enterprise and finance | Decision-making and strategy | There is a voiced need for more informed decisions in financial institutions (e.g., credit unions), suggesting a potential role for AI and considerations for how to integrate it. | Steps for integrating AI into financial management: 1) Understanding AI and its potential impact; 2) Developing a strategic AI roadmap; 3) Building internal AI capabilities; 4) Upgrading data management infrastructures; and 5) Implementing AI in stages. | Y (Technological advances, Operational efficiencies, Functionality) | Y (Operational, Technological, Ethical) |
| Christou et al 2023 | Conference proceeding | Greece, Japan | Data science, IT and software engineering | Data management and security; Decision-making and strategy | Integrating AI, Internet of Things (IoT) and edge computing offers an opportunity to enhance connectivity and develop smart, autonomous systems. This convergence can optimise operations and human-machine interaction by providing advanced data management capabilities. | Artificial Intelligence of Things (AIoT) involves the convergence of IoT and AI, constructing infrastructures with cognitive behaviour and decision-making capabilities, requiring minimal human interaction. Implementing AIoT involves data collection and pre-processing, model selection, training and optimisation, model deployment, real-time prediction, and monitoring and feedback. | Y (Technological advances, Operational efficiencies, User focus, Functionality) | Y (User, Operational, Technological,) |
| Lin 2023 | Perspective | USA | Business, enterprise and finance; Data science, IT and software engineering (e.g., Granica) | Data management and security | Interest in large language models (LLMs) has heightened focus on data management and security. Training LLMs however requires ready access to vast amounts of data, making its storage, processing and protection costly for any company. | Syneos Health has implemented a comprehensive data management strategy involving the development of feature stores and data cleaning initiatives to support AI model training, while startups like Granica offer solutions to reduce cloud storage costs and enhance cybersecurity for companies using LLMs. | Y (Technological advances, Operational efficiencies) | Y (User, Operational, Technological, Ethical) |
| Chance 2023 | Magazine | USA | Research and industry | Data management and security; Operations management and maintenance | In the digitised world, there is a push towards automation, particularly in supply chain operations. However, achieving this hinges on ensuring data interoperability and machine comprehensibility at scale. | Knowledge graphs leverage diverse data into semantic ontologies, enabling enterprises to automate workflows and answer complex questions. Implemented at scale, this infrastructure supports connectivity, enhances AI implementations and promotes a more intuitive, automated modelling approach that mirrors human thought processes. | Y (Technological advances, Operational efficiencies, User focus, Functionality) | N |
| Miguel et al 2023 | Preprint | USA | Data science, IT and software engineering | Data management and security; Data authentication; Data analytics and visualisation | Effective data management and high-quality data standards are crucial for data applications such as AI. | Semantic web standards leverage unique identifiers and metadata annotation to enable compliance with FAIR data principles (findability, accessibility, interoperability, reusability). The Research Description Framework serves a framework for describing data assets and their business context, facilitating queries and retrieval of knowledge to support diverse data governance use cases. | Y (Technological advances, Operational efficiencies, User focus, Functionality) | Y (User, Operational, Technological) |
| Engel et al 2022 | Journal article | USA, Spain | Business, enterprise and finance; Data Science, IT and software engineering (e.g., IBM Research) | Operations management and maintenance | Organisations’ service chains require integrated key performance indicators (KPIs) for operational efficiency, yet the unique service level agreements (SLAs) they entail pose challenges in aligning AI-driven decision-making with standardised monitoring systems. | AI-supported SLA analytics can be integrated into core workflows to manage complex service chains across industry sectors and public administrations. These AI-enabled service chains leverage data-intensive AI technologies to optimise decision-making processes influenced by diverse operational factors. | Y (Technological advances, Operational efficiencies, Functionality) | Y (User, Operational, Technological) |
| Jankovic and Curovic 2023 | Journal article | Serbia | Business, enterprise and finance | Data management and security; Data analytics and visualisation | The integration of AI into sustainability strategies across European companies is raising complex environmental, social and economic challenges, and also reshaping sustainability reporting and disclosure practices across sectors. | An analysis of 240 Serbian companies, using the AI Adoption Index, revealed insights regarding different levels of AI strategic planning, and the importance of effective data management, using metrics such as ‘years using big data/AI’, ‘share of processes involving big data/AI’ and ‘total AI applications’. | Y (Operational efficiencies) | N |
| Chache et al 2022 | Journal article | Russia | Public administration, policy and legislature (European Commission, UNESCO) | Decision-making and strategy | In 2021 the 41^st^ UNESCO session saw 193 countries adopting the first-ever global recommendation on AI ethics, comprising 141 items. This aimed to establish a universal framework of values, principles and actions to guide countries in shaping their AI legislations. | Key AI principles include proportionality, safety, fairness, sustainability, privacy, human oversight, transparency, responsibility, awareness and multi-stakeholder collaboration. | Y (User focus, Functionality) | Y (Technological, Ethical) |
| Ionescu and Diaconita 2023 | Review | Romania | Data science, IT and software engineering (e.g., Amazon) | Data management and security; Data analytics and visualisation; Decision-making and strategy | The current digital era highlights a significant shift towards data-driven decision-making in finance, presenting promising opportunities for data management and analysis while also introducing challenges in handling large data volumes. | Cloud service providers like AWS, Azure and GCP enhance the financial sector’s data management and analytics by offering scalable, flexible solutions for handling large volumes of data, seamless integration and advanced big data capabilities, enabling real-time decision-making, improved operational efficiency, and superior client experiences. | Y (Technological advances, Operational efficiencies, User focus, Functionality) | Y (Operational, Technological) |
| Tawil et al 2022 | Preprint | UK | Business, enterprise and finance | Data management and security; Data analytics and visualisation | Digitalisation is crucial for small to midsize enterprises (SMEs) transitioning to online and hybrid operations (e.g., post-Covid), but challenges such as low awareness of data value, absence of sector-specific data, limited resources and outdated knowledge hinder effective business transformation and AI adoption | Digitalisation plans should involve identifying key performance indicators, data collection, visualisation and interpretation. SMEs should also work on raising awareness, training staff, building a datacentric culture, address financial gaps and increase capacity for in-house data solutions (or foster collaboration). | Y (Technological advances, Operational efficiencies, User focus, Functionality) | N |
| Kelley 2022 | PhD thesis | Canada | Business, enterprise and finance | Data management and security; Compliance and risk assessment | Increasing reports of financial institutions using AI unethically highlight the importance of addressing discrimination, privacy breaches, and uninformed decision-making, while focusing on communication, management support and training, ethics and reporting mechanisms, organisational structure and interdisciplinary approaches to ethical AI adoption. | Employees identify the key components to effective adoption of ethical AI business practices as 1) communication, 2) Management support, 3) Training, 4) Ethics office, 5) Reporting mechanism, 6) Enforcement, 7) Measurement, 8) Accompanying technical processes, 9) Sufficient technical infrastructure, 10) Organisational structure, and 11) an Interdisciplinary approach. | Y (Technological advances, Operational efficiencies, Functionality) | Y (User, Operational, Technological, Ethical) |
| Korolov 2023 | Perspective | USA | Data science, IT and software engineering | Compliance and risk assessment | Companies face challenges in deploying generative AI (genAI) due to compliance and cost issues with cloud-based platforms (e.g., OpenAI), as well as the need for technical expertise in local models and risk management (e.g., to protect personal data). | VMware Private AI Foundation with Nvidia is a fully integrated genAI platform that enables companies to run AI models (e.g., Llama 2) on premises/privately and addressed compliance, cost and data privacy issues while simplifying deployment with pre-packaged models and training frameworks. | Y (Technological advances, Operational efficiencies, User focus, Functionality) | Y (Technological) |
| Jiang et al 2022 | Journal article | China | Research and industry | Operations management and maintenance | AI-enabled software and hardware are transforming manufacturing firms’ innovation strategies, however the potential benefits are tempered by risks such as bias, security breaches and privacy violations – all of which can reduce stakeholder trust and disrupt product development. | Organisations, and especially small to midsize enterprises, should assess how they manage AI applications and use AI-driven platforms to facilitate stakeholder interactions at different stages of product development in order to continuously innovate in rapidly changing environments. | Y (Technological advances, Operational efficiencies, Functionality) | Y (User, Operational, Technological) |
| Currie 2023 | Perspective | Australia | Healthcare | Research and insights; Data analytics and visualisation | There is debate around the benefits of AI tools like ChatGPT for research and problem-solving vs the risks of misuse in the health sector, urging the need for ethical guidelines on using these tools in research and education. | ChatGPT has exciting applications in medical education, research and the clinic (e.g., fact-checking, improving writing, assisting in data analysis), but its potential for errors or fabricating information poses significant risks to ethics, integrity and professionalism, leading to policies against its use (e.g., in academic publishing). | Y (Technological advances, Operational efficiencies, User focus, Functionality) | Y (User, Operational, Technological, Ethical) |
| França 2023 | Perspective | Brazil | Research and industry; Data science, IT and software engineering (e.g., Google) | Data analytics and visualisation; Research and insights | Availability of AI research tools is growing, but scientists must be aware of the options, which to choose for what purpose, and understand the rationale behind the choices they make. | AI tools like Google Scholar, Iris.ai, and Elicit.org help scientists find relevant literature, while platforms like ChatPDF, ChatGPT, and MidJourney aid in academic content analysis. Other tools (e.g., Whisper API) transcribe and describe multimedia data and support qualitative analysis (Atlas.TI's AI-driven coding assistant). | Y (Technological advances, Operational efficiencies, User focus, Functionality) | Y (User, Operational, Technological, Ethical) |
| Milmo 2023 | Magazine | UK | Data science, IT and software engineering (e.g., OpenAI) | Decision-making and strategy; Data management and security | Companies developing powerful AI systems must include independent board members representing societal interests to ensure their democratic governance and prevent misuse. | The new-look OpenAI board serves as an example where inclusion of independent board members has helped raise concerns over public AI safety, ranging from mass-produced disinformation to biased outcomes and tech-washing. | N | Y (User, Operational, Technological, Ethical) |
| Kusal et al 2022 | Review | India, UK, USA, Russia | Other (generic) | Administration and communications | Technological advancements have narrowed the communication gap between humans and machines, with research into conversational agents garnering growing interest over the years. However, these human-machine interactions also raise issues that cannot be addressed due to current technological limitations. | Conversational AI (also known as chatbots, virtual assistants) are enabled by fusion of machine learning, deep learning and natural language processing, and have versatile applications across industries. With the ability to process human input through gestures, speech or text, these agents can support administrative activities and communication (e.g., customer support), save costs and enhance customer satisfaction. | Y (Technological advances, Operational efficiencies, User focus, Functionality) | Y (User, Operational, Technological, Ethical) |
| Sheikh, Prins and Schrijvers 2023 | Book | Netherlands | Public administration, policy and legislature (Netherlands Scientific Council for Government Policy) | Decision-making and strategy | Europe's proposed AI Act signals the need for tailored regulations, yet gaps remain in existing frameworks which may require amendments to accommodate these advancements, reflecting ongoing commentary and debate over the past years. | There are concerns about the broad impact of AI, the legal and regulatory requirements necessary for AI research and development, whether rules should be tailored (e.g., case by case), and how to achieve AI transparency and explainability (e.g., through specific regulatory frameworks rather). | N | Y (User, Operational, Technological, Ethical) |
| Kanbach et al 2023 | Journal article | Germany, India | Research and industry | Data analytics and visualisation | There are discussions around the business implications of generative AI (genAI), integrating AI with business model innovation (BMI), and the role of AI and analytics in management contexts. There is also interest in how firms adapt and innovate in response to external factors like technological progress and AI. | genAI enables the creation of diverse content types such as text, images, audio, code, and videos, presenting significant implications for businesses, particularly in terms of business model innovation (BMI). These include impact on innovation activities, the work environment, and information infrastructure and involve the skills and creative thinking of people, job roles, and resources. | Y (Technological advances, Operational efficiencies, User focus, Functionality) | N |
| Holm et al 2022 | Report | UK, Norway | Research and industry (Research Council of Norway, Research on Research Institute) | Data management and security; Data analytics and visualisation; Decision-making and strategy | The Research Council of Norway and UK’s Research on Research Institute hosted a workshop for funders to share insights, discuss responsible AI/ML use in research management, and explore the adaptation of pre-trained algorithms for research. | Research funders use natural language processing on the data they collect (e.g., publications, administrative records) to enhance efficiency in funding proposal analysis and selection, as well as monitoring funded awards, however its application in strategic analysis and impact assessments remains less developed. | Y (Technological advances, Operational efficiencies, User focus, Functionality) | Y (User, Operational, Technological) |
| International Science Council 2023 | Report | France | Research and industry; Public administration, policy and legislature (e.g., OECD) | Data management and security; Decision-making and strategy | There is an ontological gap between AI principles and their integration into regulatory and governance frameworks, requiring a systems approach that includes the scientific community and considers the broad implications of AI for science and society. Effective regulation will necessitate risk-informed decision-making across multiple layers and stakeholders. | An adaptive analytical framework (derived from the OECD AI Classification Framework and INGSA’s digital wellbeing report) is proposed to guide discourse and decision-making for stakeholders using a comprehensive checklist addressing multiple layers of AI impact (from individual to economic) and encompassing short and long-term considerations. | Y (Technological advances, Operational efficiencies, User focus, Functionality) | Y (User, Operational, Technological, Ethical) |
| Rodgers, Ellingson and Chatterjee 2023 | Perspective | UK, USA, Argentina, Uruguay | Research and industry (F1000Research) | Data management and security | The European Commission emphasises the importance of open data and data sharing for AI/ML research due to the substantial volumes of data needed for model training. The issue is emphasised by the rise of AI work, which often uses existing artwork without the artists’ permission, indicating a need for freely shared data and modern copyright legislation to protect intellectual property. | Open publication and peer review are vital for responsible AI use. F1000Research model promotes an open research culture that aligns with funder mandates and supports AI researchers needing ample data for machine learning. Sharing research and data openly ensures users understand the underlying work, helps legal experts and policymakers stay updated, and allows AI researchers to build on prior developments. | Y (User focus) | Y (Ethical) |
| Department for Science Innovation and Technology 2024 | Report | UK | Public administration, policy and legislature | Decision-making and strategy | A UK gov white paper proposed a regulatory framework for AI to ensure responsible innovation, setting five principles for regulators to apply within their domains. This suggested a central government function for risk assessment and regulatory coordination. | The paper proposed 5 principles for regulators: 1) safety, security and robustness; 2) transparency and explainability; 3) fairness; 4) accountability and governance; and 5) contestability and redress. It includes supporting regulatory capability and coordination, addressing specific risks, preparing UK workers for an AI-enabled economy, enabling IP protection, and protecting citizens from AI bias. | Y (Technological advances, Functionality) | Y (User, Operational, Technological, Ethical) |
| Born 2024 | Magazine | USA | Research and industry | Decision-making and strategy | OpenAI's surprise release of GPT-4 has left some funders unprepared for the immediate risks and vast opportunities presented by generative AI. There is a need to support groups working on public interest technology, cyber policy, and responsible technology to bolster their capacity in addressing these emerging challenges effectively. | Initiatives like the NIST AI Risk Management Framework, Partnership for Public Service's leadership training, OpenMined's transparency infrastructure, and the proposed NAIRR bill exemplify efforts to enhance AI governance, transparency, and societal impact assessment globally. These focus on AI government capacity and use, greater collaboration and advocacy in AI funding, best practices and public interests, and changes around AI public narrative and legal theory. | Y (Technological advances, Functionality) | Y (User, Operational, Technological, Ethical) |
| The Standing Together Collaboration 2023 | Report | UK | Research and industry (e.g., National Institute for Health and Care Research) | Data management and security | The development of AI health technologies relies on careful selection and appropriate use of datasets to enhance health and wellbeing outcomes. | Research organisations must ensure datasets for AI health technologies are well-documented for traceability and auditability, identify groups at risk of disparate performance or harm in advance, justify appropriate dataset use for intended populations, evaluate performance across identified groups, report limitations or misuse and address uncertainties/risks with mitigation plans throughout the AI health technology lifecycle. | N | Y (User, Operational, Technological, Ethical) |
| Chubb, Cowling and Reed 2022 | Journal article | UK | Research and industry (higher education institutions, publishers) | Data analytics and visualisation; Research and insights | There is growing concern about careers, knowledge and norms in academia. Discussions on how AI affects productivity are abundant, however evidence on the impact of AI and digital technologies on research and science culture is limited. At the same time, there is increasing interest in AI tools that support various facets of academic life beyond productivity and the future of work debates. | Publishers and research institutions have begun integrating AI tools for various tasks such as reviewer selection, summarising findings, statistical analysis and literature searches; however, concerns regarding bias in AI tools and their impact on academic labour and integrity remain, highlighting the need to balance gains in productivity with potential ethical (and personal) risks. | Y (Technological advances, Operational efficiencies, User focus, Functionality) | Y (User, Ethical) |
| The Equality Trust 2024 | Report | UK | Research and industry; Healthcare | Data management and security; Data analytics and visualisation; Decision-making and strategy | Integrating AI/ML in cardiovascular healthcare is complex, highlighting potential benefits alongside risks that could exacerbate health disparities. Central to effective AI implementation is the use of comprehensive, reliable, and unbiased data, which currently lacks robust representation across all demographic groups – posing risks of health inequality. | There is a need for an intersectional AI system, mitigating bias in data interpretation, and developing health inequalities policies concurrently with AI policies, with emphasis on personalised patient management, minimising diagnostic errors, and analysing medical images to detect patterns and anomalies in cardiovascular health. | Y (Technological advances, Operational efficiencies, User focus, Functionality) | Y (User, Operational, Technological, Ethical) |
