## Supplementary material for "Exploring the potential benefits and challenges of artificial intelligence for research funding organisations: a scoping review": S3 Appendix abbreviations

ACAI	Accelerated Cloud for Artificial Intelligence

AHRC	Arts and Humanities Research Council

AI	Artificial Intelligence

AIDE Project	Artificial Intelligence in Healthcare for All

AIMDP	Artificial Intelligence Modern Data Platform

AMRC	Advanced Manufacturing Research Centre

ARMA	Association of Research Managers and Administrators

AMS	Academy of Medical Sciences

AWS	Amazon Web Services

BEIS	The Department of Business, Energy and Industrial Strategy

CDP	Cloudera Data Platform

CIHR	Canadian Institutes of Health Research

CRUK	Cancer Research UK

DBTA	Database Trends and Applications

DLHub	Data and Learning Hub for Science

DSIT	Department for Science, Innovation and Technology

EC	European Commission

EOAR	AI ethics orchestration and automated response

ESG	Environmental social and governance

ESRC	Economic and Social Research Council

EPSRC	Engineering and Physical Sciences Research Council

ETL	Extract transform and load

EU	European Union

FaaS	Function as a Service

FAIR	Findable Accessible Interoperable and Reusable

FAIR4HEP	Findable Accessible Interoperable and Reusable high-energy physics

GDPR	General Data Protection Regulation

GCP	Google Cloud Platform

GPAI	The Global Partnership on Artificial Intelligence

GPT	Generative Pre-trained Transformer

HPC-FAIR	High Performance Computing Findable Accessible Interoperable and Reusable

ICSA	International Conference on Software Architecture

ICT	Information and communication technology

IEEE	The Institute of Electrical and Electronics Engineers

INGSA	International Network for Governmental Science Advice’s

IPPH	Institute of Population and Public Health

IoT	Internet of Things

IT	Information Technoology

JBI	Joanna Briggs Institute

LLM	Large Language Models

MIT	Massachusetts Institute of Technology

MDF	Materials Data Facility

ML	Machine Learning

NIH	National Institutes of Health

NIHR ARC	National Institute for Health and Care Research Applied Research Collaboration

NIHR CRN	National Institute for Health Care Research Clinical Research Network

NIHR	National Institute for Health and Care Research (NIHR)

NLP	Natural Language Processing or Neuro-Linguistic Programming

OECD	The Organization for Economic Cooperation and Development

PRISMA	Preferred Reporting Items for Systematic reviews and Meta-Analyses

PRISMA-ScR	Preferred Reporting Items for Systematic Reviews and Meta- analysis extension for scoping reviews

R&D	Research & Development

RDF	Research Description Framework

RFO	Research Funding Organisations

RoRI	Research on Research Institute

R&T	Responsible and trustworthy

SNL	Sandia National Laboratories

SME’s	Small to midsize enterprises

UKRI	UK Research and Innovation

UNESCO	United Nations Educational, Scientific and Cultural Organization

US	United States

XAI	Explainable Artificial Intelligence
