## Supplementary material for "Exploring the potential benefits and challenges of artificial intelligence for research funding organisations: a scoping review": S3 Table key organisations

**Table S3: Key research funding and professional organisations on AI: announcements and activities**

The table below describes the priorities and positions of key funding and professional organisations in research with regards to funding developments in big data and AI, including the use of AI in research. Thirteen organisations were reviewed (listed alphabetically) for announcements or activities relevant to AI. Links to web pages, brief descriptions of page contents, and the dates the web pages were accessed, are included in the table below.

| Funding or Professional Organisation | Announcement or activity | Description and AI relevance | Date(s) accessed |
| --- | --- | --- | --- |
| Academy of Medical Sciences | [A joint statement on the AI Safety Summit (2023)](https://acmedsci.ac.uk/more/news/a-joint-statement-on-the-ai-safety-summit) | The Academy of Medical Sciences, British Academy, Royal Academy of Engineering and Royal Society have released a joint statement after the conclusion of the Artificial Intelligence (AI) Safety Summit 2023. Mentioned was the need for a collaborative, interdisciplinary approach to define how AI is governed and can be developed, deployed, and used safely and ethically – including experts from the medical, engineering, humanities, and social sciences. | 31 Jan 2024 |
| Association of Medical Research Charities | [AMRC’s response to NHS ‘Artificial Intelligence: How to get it right’ report (2019)](https://www.amrc.org.uk/News/amrcs-response-to-nhsx-artificial-intelligence-how-to-get-it-right-report)  [Annual award announced for medical research charities wishing to benefit from AI (2017)](https://www.amrc.org.uk/News/annual-award-announced-for-medical-research-charities-wishing-to-benefit-from-artificial-intelligence) | AMRC welcomed the NHSX report, stating it was helpful and comprehensive in setting out where technology can be used in the NHS and the policy work required to ensure safety, effectiveness, and ethics. Also highlighted was the need for collaboration and partnership across NHS and wider stakeholders in recognising AI’s true potential, and that the patient and public benefit is maximised.  Benevolent AI, one of the world’s largest private AI companies, has partnered with AMRC to create the new BenevolentAI award. This will be given to a single applicant once a year, and will help accelerate new treatment and therapies using AI (e.g., for analysing vast quantities of data and drug discovery) | 31 Jan 2024 |
| Association of Research Managers and Administrators | [Research Offices Survey 2020](https://zenodo.org/records/3935851) | Analysis and insights from ARMA’s first national survey of research offices across 54 UK Higher Education Institutions (HEIs). This identified AI and big data bring as the next big change to research and research management and administration practices beyond 2020. | 31 Jan 2024 |
| Canadian Institutes of Health Research | [Transforming Public Health: IPPH Strategic Plan, 2022-2026](https://cihr-irsc.gc.ca/e/53051.html)  [CIHR is seeking members for its new Advisory Committee on Ethics (2023)](https://cihr-irsc.gc.ca/e/53605.html)  [AI for Public Health Equity – Workshop Report (2019)](https://cihr-irsc.gc.ca/e/51425.html)  [CPHA Collaborator Panel Session: Exploring the Ethics of AI Approaches in Public Health (2018)](https://cihr-irsc.gc.ca/e/51249.html) | Equitable AI and equipping public health with new methods to advance health equality is a priority area in the strategic plan agreed between by CIHR’s Institute of Population & Public Health and Canada’s Chief Public Health Officer.  CIHR is seeking members with expertise and/or experience in AI for the new Advisory Committee on Ethics.  The report from the 2019 workshop in Toronto lists key themes and recommendations on maximising inclusion in data used for research, preventing and mitigating biases using AI, using AI to promote public health equity, and encouraging interdisciplinary research and collaboration, among other issues.  Key messages and reflections from a panel discussion at Public Health 2018 in Montreal, hosted by the CIHR’s Institute of Population and Public Health in collaboration with the Canadian Institute for Advanced Research, on the ethical challenges of integrating AI approaches into public health research and practice. Among numerous critical issues, these raise the importance of fairness, accountability, and transparency in AI. | 4 Apr 2024 |
| Cancer Research UK | [Policy on the use of generative artificial intelligence tools in Cancer Research UK funding applications (2023)](https://www.cancerresearchuk.org/funding-for-researchers/applying-for-funding/policies-that-affect-your-grant/policy-on-the-use-of-generative-ai-tools-in-cancer-research-uk-funding-applications?_gl=1*1vnsx87*_ga*OTA4MjM1Nzk5LjE3MDY3MDgyMDg.*_ga_58736Z2GNN*MTcwNjcwODIwOC4xLjEuMTcwNjcwODIxNS4wLjAuMA..*_gcl_aw*R0NMLjE3MDY3MDgyMDguQ2p3S0NBaUFfT2V0QmhBdEVpd0FQVGVRWi1aemJZMGo4bV9hXzNsUnRiWHQxNEs5YmRvcWt5YUg3cHZNeG9UdnFpQkpPRGRUcnFVZl9Sb0NOYUFRQXZEX0J3RQ..*_gcl_dc*R0NMLjE3MDY3MDgyMDguQ2p3S0NBaUFfT2V0QmhBdEVpd0FQVGVRWi1aemJZMGo4bV9hXzNsUnRiWHQxNEs5YmRvcWt5YUg3cHZNeG9UdnFpQkpPRGRUcnFVZl9Sb0NOYUFRQXZEX0J3RQ..*_gcl_au*MjczMTc5MzI5LjE3MDY3MDgyMDg.)  [Detect cancer earlier by interrogating medical and non-medical data sets using machine and deep learning](https://www.cancerresearchuk.org/funding-for-researchers/cancer-grand-challenges/artificial-intelligence?_gl=1*1vnsx87*_ga*OTA4MjM1Nzk5LjE3MDY3MDgyMDg.*_ga_58736Z2GNN*MTcwNjcwODIwOC4xLjEuMTcwNjcwODIxNS4wLjAuMA..*_gcl_aw*R0NMLjE3MDY3MDgyMDguQ2p3S0NBaUFfT2V0QmhBdEVpd0FQVGVRWi1aemJZMGo4bV9hXzNsUnRiWHQxNEs5YmRvcWt5YUg3cHZNeG9UdnFpQkpPRGRUcnFVZl9Sb0NOYUFRQXZEX0J3RQ..*_gcl_dc*R0NMLjE3MDY3MDgyMDguQ2p3S0NBaUFfT2V0QmhBdEVpd0FQVGVRWi1aemJZMGo4bV9hXzNsUnRiWHQxNEs5YmRvcWt5YUg3cHZNeG9UdnFpQkpPRGRUcnFVZl9Sb0NOYUFRQXZEX0J3RQ..*_gcl_au*MjczMTc5MzI5LjE3MDY3MDgyMDg.#details60)  [Unleashing the power of data to beat cancer: our research data strategy (2022)](https://www.cancerresearchuk.org/sites/default/files/cancer_research_uk_-_research_data_strategy.pdf) | The policy sets out CRUK’s position on the use of AI tools in CRUK funding applications and includes definitions, requirements for applicants and peer reviewers, and actions the consequences of breaching requirements.  Announcement of the Cancer Grand Challenges global funding platform (£20 million) for international researchers to develop innovative ways of interrogating medical and non-medical data for earlier cancer detection, including exploring pattern recognition algorithms and developing machine learning approaches to optimise algorithms during collection of anonymised datasets.  CRUK’s Research Data Strategy involves harnessing big data and AI to glean insights from large-scale health-relevant data leading to discoveries and earlier detection and treatment in cancer. Emphasised is the plan to foster a data culture that best serves the research community and drawing more on the expertise of data scientists and AI specialists. | 31 Jan 2024 |
| European Commission | [Excellence and trust in artificial intelligence (the AI Act 2023)](https://commission.europa.eu/strategy-and-policy/priorities-2019-2024/europe-fit-digital-age/excellence-and-trust-artificial-intelligence_en)  [A Europe fit for the digital age](https://commission.europa.eu/strategy-and-policy/priorities-2019-2024/europe-fit-digital-age_en)  [AI Watch](https://ai-watch.ec.europa.eu/index_en) | The EU’s stance on AI and actions to boost excellence and trustworthiness in AI systems for citizens, businesses and governments, including a regulatory framework based on human rights and fundamental values (the AI Act).  The EU’s [AI Act](https://digital-strategy.ec.europa.eu/en/policies/regulatory-framework-ai#:~:text=The%20AI%20Act%20introduces%20specific,to%20continue%20or%20step%20back) is the first-ever legal framework on AI worldwide and intends to ensure that the AI systems used across EU are safe, transparent, ethical, unbiased and under human control. This introduces categories of risks for AI systems (from unacceptable to minimal), and rules for providers of high-risk systems. In 2018, the EC and EU Member States also developed a (updated in 2021) Coordinated Plan on AI to lay the ground for national strategies and policy developments. The four key policy objectives are 1) setting enabling conditions for AI’s development and uptake, 2) building strategic leadership in high-impact sectors, 3) making the EU the right place for AI to thrive, and 4) ensuring AI technologies work for people.  As part of the above efforts, the EC will also be setting up R&I infrastructure and funding a variety of AI projects.  The EU’s digital strategy aims to facilitate the digital transformation of businesses, empower people with a new generation of technologies, and achieve a target of a climate-neutral EU by 2050. The strategy involves [establishing standards in data](https://op.europa.eu/en/publication-detail/-/publication/b11a0504-75eb-11ed-9887-01aa75ed71a1/language-en), improving digital skills, and investing in the necessary digital technology and infrastructure, including AI (above).  The web page lists projects the EU is working on, including through the EC, and strategy timeline updates.  The EC are monitoring the development, uptake, and impact of AI for Europe via access to country AI strategy reports, AI landscape, and investment dashboards (via the web pages interactive AI in Europe map tool). The page also lists relevant events, news, and European AI sites. |  |
| Gates Foundation | [The first principles guiding our work with AI (2023)](https://www.gatesfoundation.org/ideas/articles/artificial-intelligence-ai-development-principles)  [AI equity: Ensuring access to AI for all](https://www.gatesfoundation.org/ideas/science-innovation-technology/artificial-intelligence) | The Gates Foundation is interested in making AI technology broadly available societies and economies, and transformative to how people communicate, work, learn and improve their wellbeing. Mitigating uneven benefits from AI (e.g., due to lack of access) by ensuring beneficiaries participate in the development technology is a particularly important goal. With this focus on access and equity in AI work, the foundation therefore established an internal Global AI Task Force in March 2023 to help define the role of AI, establish foundation principles with regards to AI and coordinate further activities, and drive a responsible approach to the foundation’s engagement with AI usage to ensure it is safe, ethical and equitable.  A further look at the Gates Foundation’s guiding AI principles and how these drive current efforts to ensure low- and middle-income countries and included in the (locally relevant) creation of AI tools. The web page also includes links to the AI Ethics & Safety Committee, AI innovators and related articles. | 21 Mar 2024 |
| Joseph Rowntree Foundation | [AI for public good (2024)](https://www.jrf.org.uk/ai-for-public-good) | JRF is connecting people from different fields to contribute to ongoing discussions about AI and how it can be used for the public good. As part of this, JRF will be commissioning content that explores four initial areas of AI: AI narratives, AI in the public sector, AI and civil society, and AI, power, relationships, and values. | 22 Mar 2024 |
| National Institutes of Health | [The Use of Generative Artificial Intelligence Technologies is Prohibited for the NIH Peer Review Process (2023)](https://grants.nih.gov/grants/guide/notice-files/NOT-OD-23-149.html)  [Artificial Intelligence at the NIH](https://datascience.nih.gov/artificial-intelligence) | Notice to clarify on Maintaining Security and Confidentiality in NIH Peer Review: Rules, Responsibilities and Possible Consequences and inform the extramural community that the NIH prohibits NIH scientific peer reviewers from using natural language processors, large language models, or other generative AI tools for analysing and formulating peer review critiques for grant applications and R&D contract proposals.  NIH aims to make biomedical data FAIR-compliant (findable, accessible, interoperable, and reusable) and usable for AI and machine learning applications through a series of activities and initiatives (listed in the web page). These include [AI/ML Consortium to Advance Health Equity and Researcher Diversity](https://datascience.nih.gov/artificial-intelligence/aim-ahead), [Institute- and Center-Funded Initiatives](https://datascience.nih.gov/artificial-intelligence/institute-and-center-funded-initiatives) to develop and implement AI/ML technologies across biomedical research domains, and [Bridg2AI](https://www.commonfund.nih.gov/bridge2ai). |  |
| National Institute for Health and Care Research | [Data Science & AI](https://www.io.nihr.ac.uk/data-science-ai/)  [Artificial intelligence funding](https://www.nihr.ac.uk/explore-nihr/funding-programmes/ai-award.htm)  [Artificial Intelligence and Machine Learning at UCLH BRC](https://www.uclhospitals.brc.nihr.ac.uk/criu/research-impact/artificial-intelligence-and-machine-learning)  [Artificial intelligence e-learning launched for researchers (2022)](https://www.nihr.ac.uk/news/artificial-intelligence-e-learning-launched-for-researchers/31387)  [The Use of Artificial Intelligence (AI) in Systematic Reviews Masterclass 2024](https://www.arc-nt.nihr.ac.uk/news-and-events/2024/feb-24/the-use-of-artificial-intelligence-ai-in-systematic-reviews-masterclass-applications-now-open/) | The Data Science & Artificial Intelligence Programme, delivered by the NIHR Innovation Observatory (IO), is designing and developing external and internal facing intelligent data-driven tools for searching, identifying and extracting data across various open data sources for knowledge and insight. Current projects include OpenScan, a cloud-based database which uses natural language processing techniques to extract health innovation data from a variety of data sources, and the IO Toolkit to support IO activities, which includes AI-based screening tools.  The NIHR offers three funding streams to support health and care research that involves AI: the Artificial Intelligence in Health and Care Award (AI Award) that supports AI solutions across the whole development pathway; the Artificial Intelligence for Multiple Long-Term Conditions (Multimorbidity) call (AIM) for using AI and data science to address multimorbidity challenges; and the Artificial Intelligence and Racial and Ethnic Inequalities in Health and Care call that supports AI research to meet the needs of ethnic minorities.  The NIHR Biomedical Research Centre aims to transform research at University College London Hospitals by applying AI across its processes and introducing real-time operational modelling to enable prediction of individual care outcomes, causal factors in care, and potential alternative pathways to maximise care quality.  The NIHR Clinical Research Network has developed a (self-paced) AI e-learning course in partnership with Imperial College London groups to build AI awareness, knowledge and skills among clinical researchers and professionals.  NIHR’s ARC North Thames Academy is holding an AI in Systematic Reviews masterclass on 24 April 2024, aimed at nurses, allied health, social care and public health professionals, and government staff who wish to enhance their research skills through integration of AI. The course offered cutting-edge insights, practical applications, and networking opportunities. | 31 Jan and 28 Mar 2024 |
| Royal Society | [AI and data (2024)](https://royalsociety.org/current-topics/ai-data/)  [Machine learning resources](https://royalsociety.org/news-resources/resources-for-schools/machine-learning/) | The Royal Society is working on data and digital technologies to explore potential opportunities for, and barriers to, AI applications, including the implications for scientific research, society, and digital technology for the planet. The AI and data web page lists related projects and resources (e.g., see Machine learning resources below), publications, and events.  The Society’s programme of work on machine learning is investigating the potential of this technology over the next 5-10 years, with a focus on primary and secondary education and ensuring students aged 5-18 are able to develop key STEM skills for broader future career choice. As part of this programme, the Society has produced in partnership with the British Science Association a series of CREST Award resources to encourage students to explore machine learning and its capabilities. | 22 Mar 2024 |
| UK Research and Innovation | [Artificial intelligence and data economy (2024)](https://www.ukri.org/what-we-do/ukri-challenge-fund/artificial-intelligence-and-data-economy/)  [Ethics in artificial intelligence research and development (2021)](https://www.ukri.org/opportunity/including-ethics-in-artificial-intelligence-research-and-development/)  [Responsible AI UK international partnerships (2023)](https://www.ukri.org/opportunity/responsible-ai-uk-international-partnerships/)  [Responsible and trustworthy artificial intelligence (2022)](https://www.ukri.org/opportunity/responsible-and-trustworthy-artificial-intelligence/)  [AI Research Resource funding opportunity launches (2024)](https://www.ukri.org/news/ai-research-resource-funding-opportunity-launches/#:~:text=It%20follows%20the%202023%20Autumn,the%20AI%20Research%20Resource%20programme) | One of the four themes covered by the UKRI Challenge Fund (£5.6 billion) concerns AI and challenges in the data economy and aims to put the UK at the forefront of the AI and data revolution. Among the numerous goals under this funding theme, UKRI aims to support service industries to use technologies such as AI and data analytics for next generation services and invest in projects that help the UK’s digital computing infrastructure to become more secure.  The Arts and Humanities Research Council funded grants to explore ways to include ethics in the research and development of AI. These put special emphasis on networking and creating collaborations with business, third sector organisations and government bodies.  UKRI funded grants in partnership with Responsible AI UK for researchers to develop international partnerships with world-leading organisations and research centres active in establishing and promoting responsible AI practices.  The EPSRC, AHRC, ESRC and Innovate UK partnered on a £25 million award to fund and develop a leadership team to drive the UK’s responsible and trustworthy (R&T) AI agenda and build a diverse and inclusive community across disciplines and sectors.  UKRI announces a funding opportunity to support new advanced supercomputers for AI research, with support from the Department for Science, Innovation and Technology and following the Chancellor’s 2023 promise to invest £500 million into AI computing. This funding call will identify the organisations capable of hosting and operating large-scale computer systems for AI applications. | 31 Jan and 12 Mar 2024 |
| Wellcome | [Funders joint statement: use of generative AI tools in funding applications and assessment (2023)](https://wellcome.org/what-we-do/our-work/joint-statement-generative-ai)  [New programme to explore how innovation in health data can benefit everyone (2019)](https://wellcome.org/news/new-programme-explore-how-innovation-health-data-can-benefit-everyone) | The Research Funders Policy Group, of which Wellcome is a member, set out a joint statement of expectations around the use of generative AI tools in funding applications. These include researchers ensuring that these tools are used responsibly, and in accordance with relevant legal and ethical standards, and peer reviewers ensuring they keep the contents of funding application confidential when using generative AI tools to develop their application reviews.  Wellcome announced £75 million investment into a new five-year programme on data for science and health. This is focused on exploring the opportunities AI can bring to health data and removing barriers to ‘data innovation’. Specifically, through this programme, Wellcome aims to motivate more data scientists to work with health data, fund the development of open source tools, and build the public’s trust, understanding and participation in health data innovation. | 31 Jan 2024 |
